# Long-term Penetrance of Disease Variants in Genes Prioritized for Genomic Newborn Screening: Evidence from Adult Biobanks

**DOI:** 10.64898/2026.06.10.26355380

**Authors:** Nina B. Gold, Hana Zouk, Julie Yeo, Stuart Lipsitz, Satoshi Koyama, Harini Somanchi, Emma Perez, Margaret Sunitha Selvaraj, Lauren O’Grady, Emily Miller, Anna C.F. Lewis, Elizabeth W. Karlson, Alanna Strong, Jessica I. Gold, Heidi L. Rehm, Pradeep Natarajan, Robert C. Green

## Abstract

**Importance:** Genomic newborn screening (gNBS) is a potential public health intervention, but its positive predictive value (PPV) remains uncertain. Estimating the prevalence and penetrance of pathogenic and likely pathogenic (P/LP) variants in genes prioritized for screening may clarify the long-term PPV and clinical utility of gNBS.

**Objective:** To compare ICD-based ascertainment, electronic medical record (EMR) review, and clinical assessment of genetic disorders in adults with P/LP variants in 54 genes prioritized for gNBS.

**Design:** Two-cohort observational study with EMR review and clinical assessment in the hospital-based cohort.

**Setting:** The U.K. Biobank (UKB) and Mass General Brigham Biobank (MGBB).

**Participants:** 451,877 adults from the UKB and 53,371 from the MGBB, all with exome sequencing data.

**Exposures:** P/LP variants in 54 genes prioritized through expert consensus for gNBS, in genotypes consistent with each gene’s inheritance pattern.

**Main outcomes and measures:** The primary outcome was the absolute difference in the proportion of MGBB participants identified as affected by ICD versus EMR ascertainment. Secondary outcomes included findings from clinical assessments of undiagnosed MGBB participants, corrected UKB penetrance estimates, and extrapolation to U.S.. annual birth cohorts and living adults.

**Results:** P/LP variants were identified in 665 UKB participants (0.15%) and 82 MGBB participants (0.15%), approximately 1 in 650. In MGBB, EMR review revealed that 58/82 individuals (70.7%) were undiagnosed, although 25 of 58 (43.1%) had documented symptoms. Disease-associated ICD codes were found in 39.0% (32/82) of participants, whereas EMR review identified symptoms in 59.8% (49/82, McNemar P<.001). Applied to UKB, this correction yielded a penetrance of 28.4% (95% CI, 18.6% to 38.2%), implying that 73 to 203 participants beyond the 51 identified by ICD codes may have clinical features of disease. Extrapolated to U.S. birth cohorts, 4,900 to 5,700 newborns per year may harbor P/LP variants in these genes and survive into adulthood. Approximately 355,000 to 410,000 U.S. adults may have P/LP variants in these genes.

**Conclusions and relevance:** Penetrance of P/LP variants in genes prioritized for gNBS is substantially higher than ICD estimates suggest. Many adults with P/LP variants are symptomatic but undiagnosed, supporting inclusion of these genes in gNBS.

**Key points:** *Question:* What proportion of adults with genomic variants linked to treatable genetic diseases develop symptoms and what does this imply for genomic newborn screening (gNBS)?

*Findings:* Among 505,222 adults, disease-associated variants in 54 gNBS genes were found in approximately 1 in 650 participants. In a hospital biobank cohort, only 29.3% of participants had been diagnosed, but electronic medical record review and targeted phenotyping identified symptoms in 59.8%.

*Meaning:* Individuals with treatable genetic disorders that are identifiable through genomic screening are symptomatic but undiagnosed into adulthood, highlighting the importance of genomic newborn screening.

## Introduction

Genomic newborn screening (gNBS) is being evaluated internationally as an adjunct to conventional newborn screening (NBS) to identify infants at risk for treatable genetic disorders.^1–7^ Critics of gNBS often highlight that its positive predictive value (PPV), an important consideration for clinical utility, is uncertain.^8–11^

The PPV of gNBS depends upon the prevalence and penetrance of variants in the genes being screened, where penetrance refers to the probability that an individual with a pathogenic or likely pathogenic (P/LP) variant will develop features of the associated disorder.^12–14^ Because P/LP variants can be associated with a wide spectrum of clinical presentations, including mild or atypical features to no clinical disease, understanding penetrance is essential for estimating how often a genetic diagnosis predicts clinical illness.^15–17^ In many gNBS studies, infants with P/LP variants but no apparent phenotype may be classified as “false positives,” although some may later develop clinical features or have manifestations that are not recognized as part of the genetic disorder.^2,18^

Hospital– and population-based biobanks offer a window onto how variants in gNBS-prioritized genes manifest across the lifespan. Biobank data have provided insights about the prevalence and penetrance of P/LP variants associated with cancer predisposition syndromes,^19,20^ cardiac disease,^19,21,22^ inherited metabolic disorders,^23^ and multisystem conditions such as Marfan syndrome and Noonan syndrome.^24,25^ Penetrance estimates from biobanks must be interpreted carefully, however, as they are depleted of individuals with severe pediatric disorders,^24^ and the apparent absence of symptoms in participants with P/LP variants may reflect true incomplete penetrance, or other explanations including variants in *cis* configuration, X-linked disease, variant misclassification, presymptomatic disease, or limited recognition and documentation of clinical features.^13,14,25–34^ Several of these challenges apply to gNBS data as well; the immediate question is therefore not whether every mechanism underlying apparent incomplete penetrance can be resolved, but what proportion of individuals with P/LP variants identified by current genomic testing methods will develop clinical features of the associated disorder. This information may inform counseling and treatment decisions for infants with positive gNBS results.

We identified participants in the population-based U.K. Biobank (UKB) and hospital-based Mass General Brigham Biobank (MGBB) with P/LP variants in genes prioritized for gNBS, restricting analyses to genotypes consistent with each gene’s inheritance pattern (e.g., one P/LP variant associated with autosomal dominant and X-linked disorders and two P/LP variants associated with autosomal recessive disorders).^35^ To examine how ascertainment methodologies influence penetrance estimates, we compared *International Classification of Disease, 10th Revision* (ICD) codes, electronic medical records (EMR), and clinical assessments for estimating the penetrance of P/LP variants in genes prioritized for gNBS. The primary outcome was the absolute difference in the proportion of MGBB participants identified as affected by ICD versus EMR review. Secondary outcomes included corrected UKB penetrance estimates and extrapolation to United States (U.S.) and United Kingdom (U.K.) annual birth cohorts and living adults.

## Methods

### Study design

We conducted a two-cohort study using a validation-subsample design, in which the MGBB served as a deeply-phenotyped cohort with EMR review and clinical assessment, and the UKB served as a population-scale cohort with phenotypes defined by ICD codes only. This design allows the relationship between a known error-prone measurement, such as ICD ascertainment, and a gold-standard measurement, such as EMR review, to be characterized in a smaller subsample, then applied to the larger cohort.^36,37^ The Massachusetts General Hospital Institutional Review Board approved this study (protocol #2024P000954).

### Description of the UKB and MGBB

The MGBB is an EMR-linked biorepository that includes data from 53,371 participants who underwent exome sequencing and consented to be recontacted for research and clinical purposes (Table 1).^19^ UKB participants were recruited in the U.K. between 2006 and 2010 at 40 to 69 years of age (now with a median age over 70 years^38^) and are 54% female.^39^ They consented to longitudinal data collection.^40^ Exome sequencing was performed on 451,877 participants who were included in this study.^41^ Collection of demographic data in the MGBB and UKB is described in eMethods in Supplement 1.

**Table 1.** Demographic characteristics of hospital-based biobank participants. Demographic characteristics of MGBB participants who have undergone exome sequencing (n = 53,371) and those with likely pathogenic and pathogenic variants in the 54 genes included in this study (n = 82)

|  | No. of participants |  |
| --- | --- | --- |
|  | MGBB Biobank participants who underwent WES (N=53,371) | MGB Biobank participants with P/LP variants (N=82) |
| Vital Status |  |  |
| Living | 45715 | 74 |
| Deceased | 7656 | 8 |
| Gender |  |  |
| Female | 29697 | 50 |
| Male | 23672 | 32 |
| Unknown | 1 | 0 |
| Race |  |  |
| American Indian or Alaska Native | 64 | 0 |
| Asian | 1466 | 2 |
| Black | 2627 | 5 |
| Declined | 443 | 0 |
| Native Hawaiian or Other Pacific Islander | 19 | 0 |
| Other | 2215 | 2 |
| Two or More | 435 | 2 |
| Unknown/Missing | 942 | 0 |
| White | 45153 | 71 |
| Ethnicity |  |  |
| Declined | 2681 | 0 |
| Hispanic | 1594 | 5 |
| Non Hispanic | 46936 | 77 |
| Unknown/Missing | 2160 | 0 |
| Age, y |  |  |
| 0-9 | 0 | 0 |
| 10-19 | 20 | 0 |
| 20-29 | 1180 | 0 |
| 30-39 | 6495 | 16 |
| 40-49 | 6484 | 11 |
| 50-59 | 7520 | 12 |
| 60-69 | 11387 | 20 |
| 70-79 | 12146 | 16 |
| 80-89 | 6654 | 4 |
| 90+ | 1485 | 3 |

### Selection of genes and curation of genomic variants

We selected 54 monogenic disease genes associated that were recommended by ≥75% of 238 rare disease experts for inclusion in gNBS in a survey study (eTable 1 in Supplement 1).^35^ One gene, *G6PD*, was excluded due to its high prevalence and variable expressivity.^42^ Curation of P/LP variants is described in eMethods in Supplement 1.

### Phenotype ascertainment

Three ascertainment methods were applied. First, ICD ascertainment in the MGBB and UKB used a structured list of ICD codes mapped to each gene’s corresponding disorder (eTable 2 in Supplement 1). Second, structured EMR review was performed on all MGBB participants with P/LP variants using a rubric capturing symptoms, laboratory or imaging findings consistent with the condition, and documented molecular diagnoses. Third, living, undiagnosed MGBB participants were recontacted and offered a clinical visit at which additional histories, physical exam findings, and laboratory or imaging studies were sought. Ascertainment methods are described in more detail in eMethods in Supplement 1.

### Difference in ICD and EMR estimates of disease

The primary outcome was the absolute difference in the proportion of MGBB participants identified as affected by ICD-based versus EMR review-based ascertainment. The 95% confidence interval was estimated using Newcombe’s hybrid score method for paired binary proportions.^43^ McNemar’s test was reported as supporting evidence of systematic disagreement.

Corrected UKB penetrance was estimated by applying the MGBB EMR versus ICD additive correction factor to the UKB participants, for whom complete EMR data is unavailable. Next, 95% CIs were estimated using a nonparametric bootstrap with 10,000 iterations, resampling MGBB participants with P/LP variants and recalculating the absolute increase in ascertainment from ICD codes to structured EMR review. This correction was then applied to the UKB cohort to estimate the number of additional participants with potentially unrecognized clinical disease.

We repeated the primary analyses after excluding participants with *RET* c.2410G>A (p.Val804Met), a low penetrance variant that had been classified as P/LP in the source data. We also performed analyses stratified by sex for X-linked genes.

### Population extrapolation

Approximately 3.6 million infants are born in the U.S. and 660,000 in the U.K annually.^44,45^ We used these figures to estimate how many newborns each year may have P/LP variants in the genes studied and live into adulthood.

The prevalence of P/LP variants in genotypes consistent with each gene’s inheritance pattern was estimated from the pooled UKB and MGBB cohorts, and uncertainty was incorporated using the 95% CI around this prevalence estimate. Estimates were reported as approximate, order-of-magnitude projections. We applied the same approach to the living adult population, using population denominators of approximately 258 million adults in the U.S. and 53 million adults in the U.K.^45,46^

## Results

### Prevalence of P/LP variants

Among 53,371 MGBB participants (median age 61.5 years, IQR, 44.5-71.0 years), 82 (0.15%) had P/LP variants in genotypes consistent with each gene’s inheritance pattern across 10 of the 54 genes studied (Figure 1; eTable 3 in Supplement 1). In UKB, 665 participants (0.15%) had P/LP variants in 18 of the 54 genes studied (Figure 2; eTable 4 in Supplement 1).

**Figure 1.**
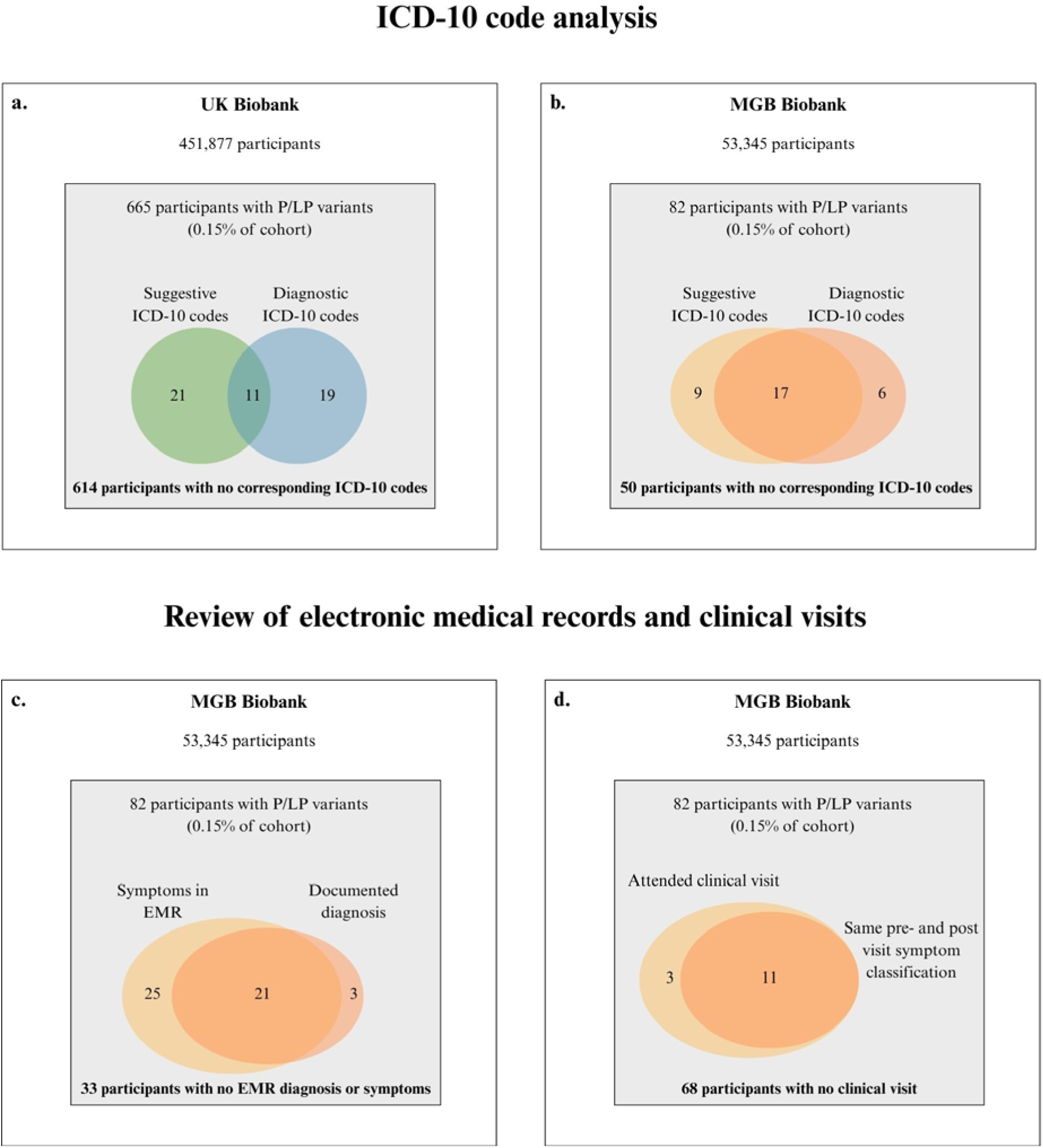
Comparison of three phenotype ascertainment methods in a hospital– and population-based biobank. ICD code analysis and electronic medical record review of participants with pathogenic/likely pathogenic variants in the UK Biobank and Mass General Brigham Biobank.

**Figure 2.**
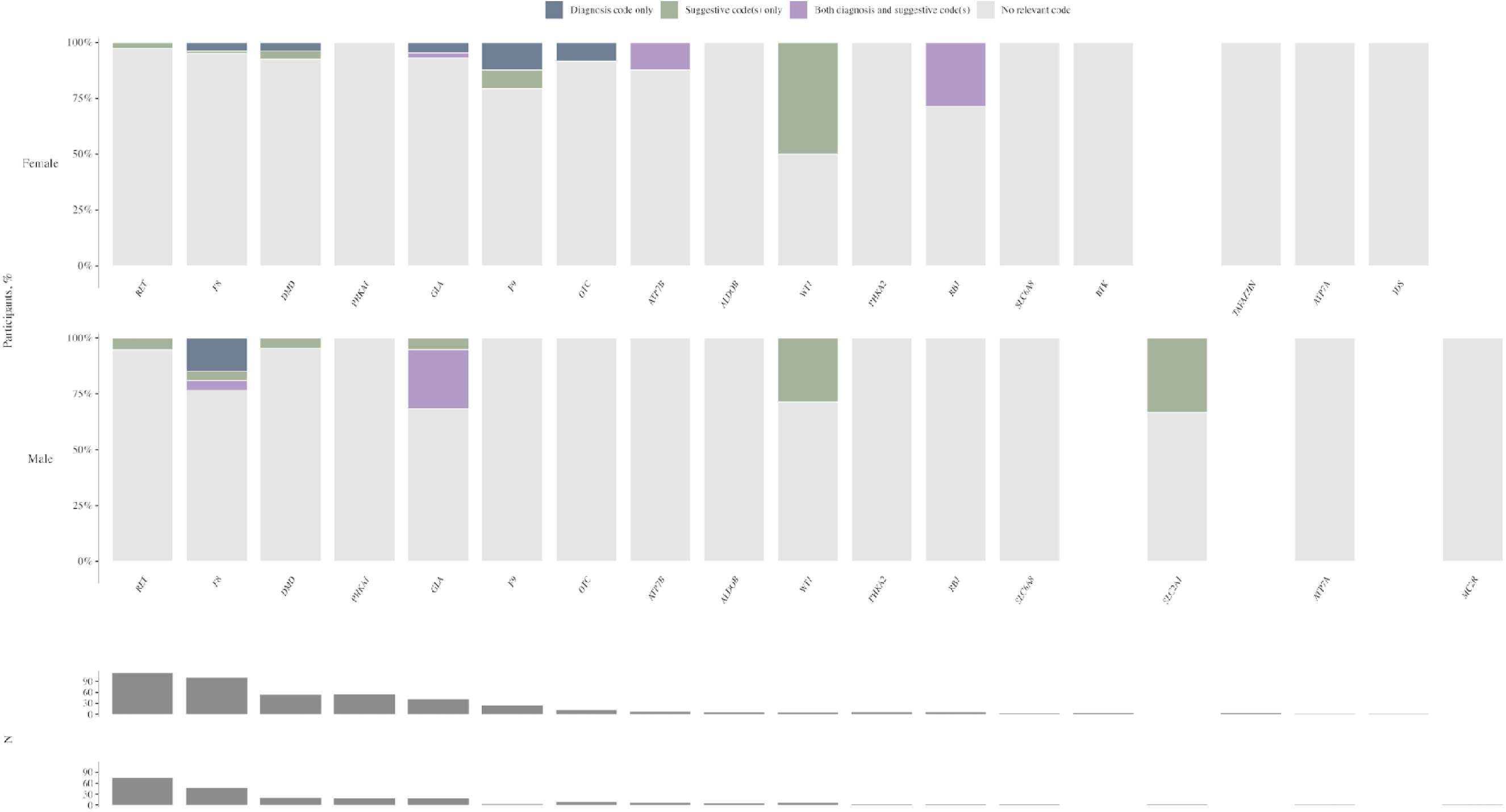
ICD code-based phenotypes identified in a population-based biobank. Participants in the U.K. Biobank with pathogenic and likely pathogenic variants in the 54 genes in this study (n = 665) and the presence of diagnostic ICD codes and ICD codes associated with highly suggestive clinical features.

In both the MGBB and UKB, P/LP variants were most common in *RET* (MGBB, n = 23, 0.04% of all participants; UKB, n = 190, 0.04% of all participants). Most participants with *RET* variants in both cohorts harbored the c.2410G>A (p.Val804Met) variant (Figure 3). In both the MGBB and UKB, *F8*, associated with *F8*-related hemophilia, was the second most common gene in which P/LP variants were identified (MGBB, n = 19, 0.04% of all participants; UKB, n = 147, 0.03% of all participants). Among participants in the MGBB and UKB with *F8* variants, 8 (42.1%) and 49 (33.3%) respectively had the c.6089G>A (p.Ser2030Asn) variant.

**Figure 3.**
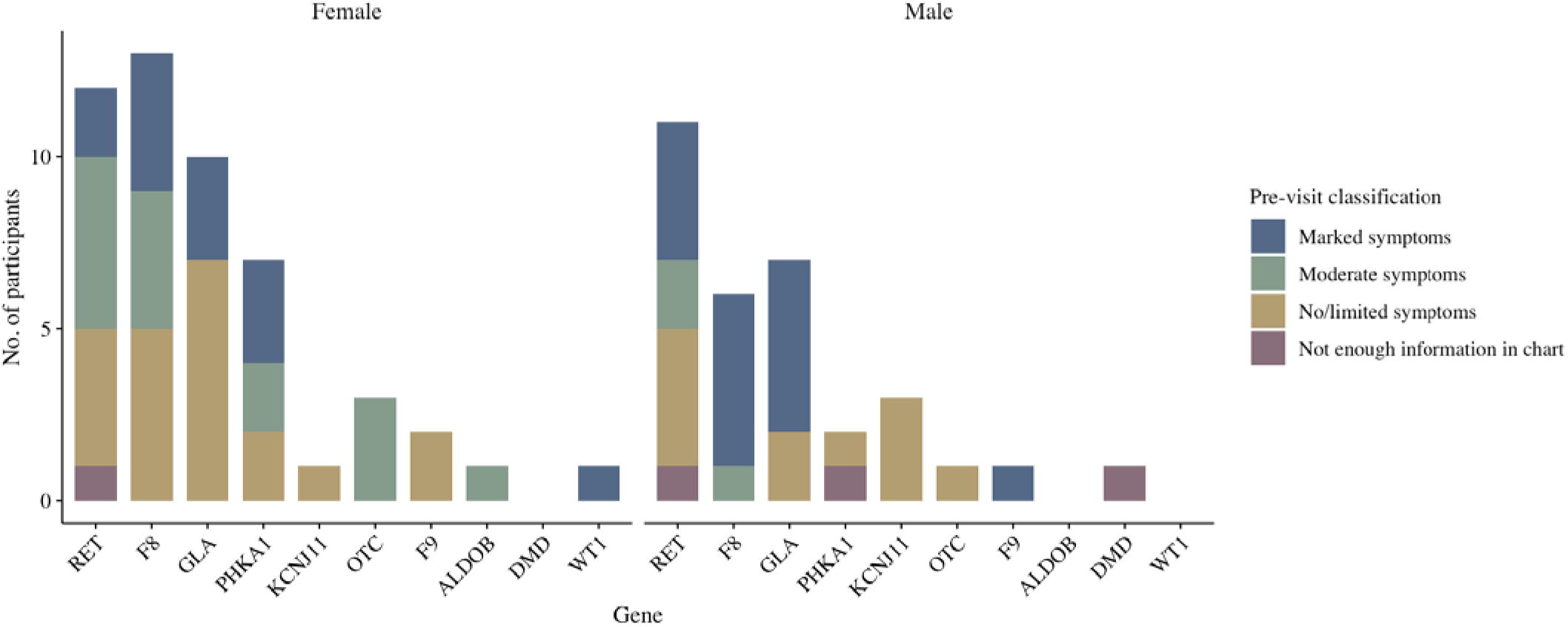
Sex-stratified analyses and symptom severity in a hospital-based biobank. Mass General Brigham Biobank participants with pathogenic and likely pathogenic variants in the 54 genes in this study (n = 82) and their symptom severity based on electronic medical record review.

### Phenotype ascertainment

Among 82 participants in the MGBB with P/LP variants, 32 (39.0%) had a diagnostic ICD code and/or an ICD code associated with suggestive symptoms of the corresponding genetic disorder. Among 665 UKB participants with LP/P variants, 51 (7.7%) had a diagnostic and/or suggestive ICD code (eTable 5 in Supplement 1; eTable 6 in Supplement 1).

In total, 49 of 82 MGBB participants (59.8%) had either a documented diagnosis or suggestive clinical manifestations recorded in the EMR. This included 21 participants with both a documented diagnosis and clinical manifestations, 25 with manifestations but no documented diagnosis, and 3 with a documented diagnosis but no recorded manifestations meeting review criteria. Among the 58 participants who did not have a documented genetic diagnosis, 15 (25.9%) had moderate symptoms and 10 (17.2%) had marked symptoms.

### Difference in ICD and EMR estimates of disease

Among the 82 MGBB participants with P/LP variants, 32 (39.0%) had ICD codes associated with the corresponding disorder and 49 (59.8%) had symptoms or a documented molecular diagnosis in the EMR, indicating an absolute increase of 20.8 percentage points of EMR over ICD ascertainment (95% CI 10.2 to 30.3 by Newcombe’s method) (eTable 7 in Supplement 1). Nineteen MGBB participants with P/LP variants had diagnoses recorded in the text of the EMR but not as ICD codes, while only 2 were ICD-positive but EMR review-negative (McNemar P<.001).

Excluding the 12 participants with *RET* p.Val804Met shifted the EMR-based penetrance estimate to 44 of 70 participants (62.9%) with clinical manifestations and/or a documented molecular diagnosis. Sex-stratified analyses for X-linked genes are shown in Figure 3.

### Clinical assessments of MGBB participants

In total, 51 of 82 MGBB participants (62.2%) were living and undiagnosed. Fourteen participants (27.5% of those eligible) completed a clinical visit at which targeted histories and testing were collected (Figure 4).

**Figure 4.**
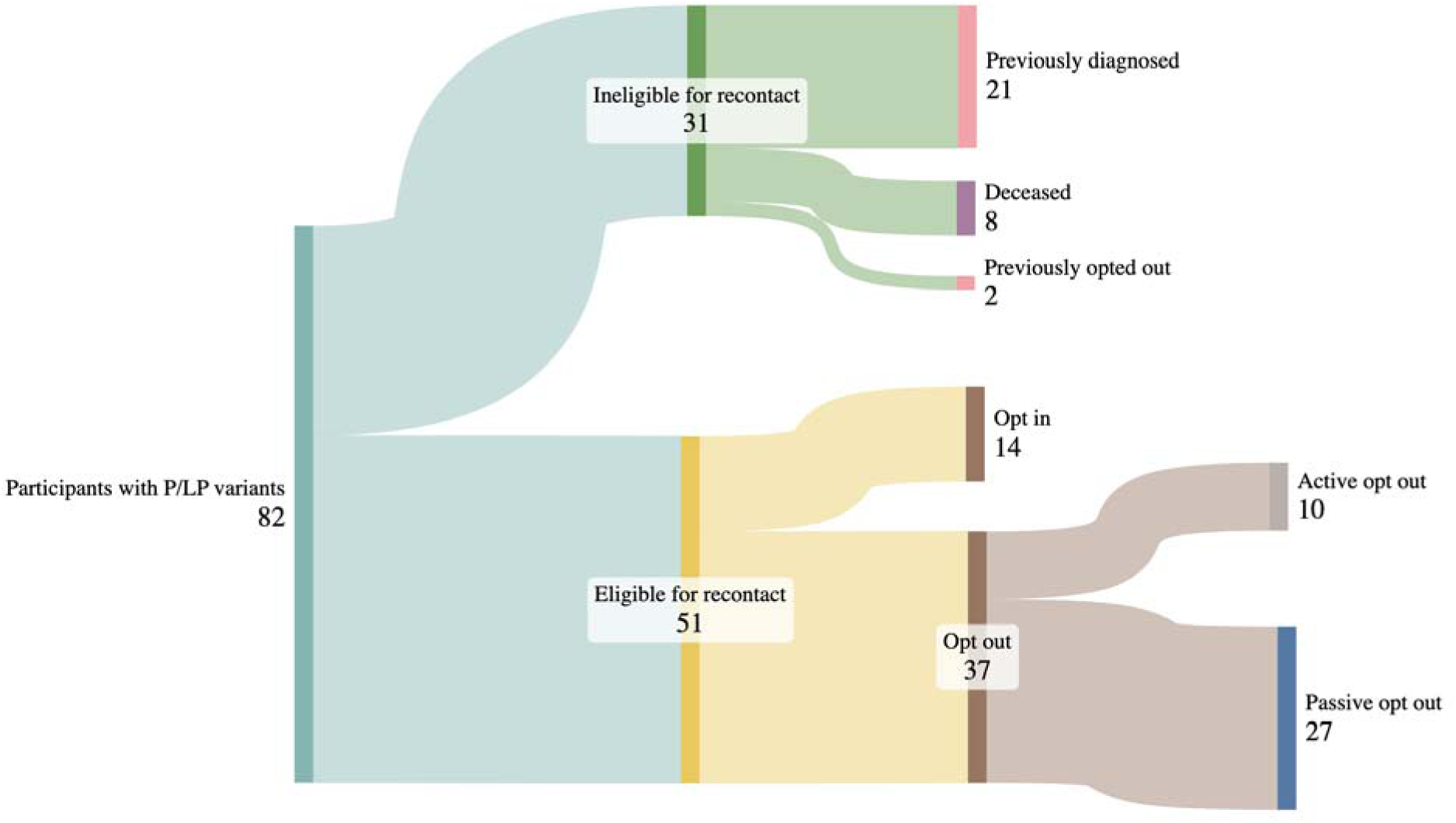
Eligibility and attendance of a clinical assessment of participants identified in a hospital-based biobank. Sankey diagram demonstrating electronic medical review, recontact, and clinical assessment of participants in the Mass General Brigham Biobank with pathogenic and likely pathogenic variants in the 54 genes in this study (n = 82).

Although visits often revealed more detailed histories, the symptom severity of most participants who attended them (11/14, 78.6%) remained unchanged. In 3 cases (21.4%), visits identified more severe manifestations of the associated disorder than were apparent from the EMR alone. For example, a female participant with a P/LP variant in *PHKA1*, associated with *PHKA1*-related glycogen storage disease, type IX, had a longstanding history of chronic muscle pain that had been dismissed by other clinicians; laboratory testing revealed an elevated baseline creatine kinase level, suggestive of metabolic myopathy. Similarly, a patient with biallelic variants in *ALDOB* had classic symptoms of *ALDOB*-related hereditary fructose intolerance, including lifelong aversion to simple carbohydrates, which had not been documented in the EMR.

### Corrected UKB penetrance

In the UKB, 51 of 665 (7.7%) participants with P/LP variants in genotypes consistent with each gene’s inheritance pattern had a diagnostic or suggestive ICD code for the corresponding disorder. Application of the additive correction yielded a corrected UKB penetrance of 28.4% (95% CI, 18.6%-38.2% by bootstrap). Under the additive correction, the implied number of UKB participants with unrecognized clinical disease beyond the 51 identified by ICD codes is approximately 73 to 203, with a point estimate of 138.

### Population extrapolation

Applied to an estimated 3.6 million U.S. live births per year, the variant prevalence (0.15%) implies that approximately 5,300 newborns per cohort carry P/LP variants in genotypes consistent with each gene’s inheritance pattern associated with disorder in one of the proposed 54 gNBS genes (95% CI interval: 4,900 to 5,700). Assuming the EMR-based penetrance of approximately 60%, at least 3,200 newborns per year would be expected to develop disease and live into adulthood, with a bootstrap interval of approximately 2,600 to 3,800. The corresponding U.K. estimate, applied to 660,000 annual live births, is approximately 470 to 700 newborns per year. Applied to a U.S. adult population of approximately 258 million, approximately 355,000 to 410,000 living U.S. adults may harbor P/LP variants in genotypes consistent with each gene’s inheritance pattern state associated with disease.

## Discussion

### Summary of findings and relevance to gNBS

gNBS is a rapidly expanding area of international research due to its potential to identify infants at risk for a wide range of treatable genetic disorders before the onset of irreversible symptoms.^8^ Several studies and reviews have highlighted prevalence and penetrance as important considerations for disease selection,^2,11,47–49^ as these factors influence both the number of infants found positive for P/LP variants and the predictive value of those findings. Because the participants in this study were adults, their data provides insights into longitudinal disease expression that gNBS studies with prospective follow-up will not be able to capture for decades.

In this two-cohort study of 505,222 adults, we found that 0.15%, or approximately 1 in 650 individuals, harbored P/LP variants in genotypes consistent with each gene’s inheritance pattern across 54 conditions prioritized for gNBS.^35^ EMR review of MGBB participants with P/LP variants identified clinical features of disease or diagnoses in 59.8%, compared with 39.0% by ICD-based ascertainment, and suggested that a high proportion of those with P/LP variants were undiagnosed. Applied to annual birth cohorts, the corrected estimate implies that several thousand U.S. newborns each year harbor such variants and would live into adulthood, many with symptoms that are unrecognized as manifestations of the underlying genetic disorder.

### Findings in the context of prior gNBS and biobank studies

These findings extend results from prior gNBS and biobank studies. Of GUARDIAN’s approximately 4,000 screened newborns, 4 (0.1%) had P/LP variants in the 54 genes included in this study, a frequency consistent with our prevalence estimate of 0.15% in adults.^2^ Low penetrance estimates based on ICD-based phenotypes have been reported in the UKB and hospital-based biobanks,^15^ and it has been well-documented previously that ICD codes are insufficient for rare diseases.^50,51^ Empirically, ICD codes for monogenic diseases often follow rather than precede clinical diagnosis^52^ and biobank penetrance estimates for several monogenic conditions rise substantially when EMR-based phenotyping is applied.^14^ The hidden burden of adults with rare monogenic disorders, particularly among patients admitted to intensive care units, has recently been estimated to be as high as 24%, most of whom were undiagnosed.^53^

The contribution of this study, therefore, is not simply the recognition that ICD codes underestimate penetrance, but a demonstration of the magnitude of that underestimation for genes prioritized for gNBS and the prevalence of undiagnosed, symptomatic adults with actionable findings, a healthcare gap that could be addressed through gNBS. When EMR data and clinical follow-up visits are incorporated, and particularly when known attenuated variants were accounted for, the estimated lifetime penetrance of P/LP variants in these genes is comparable to or exceeds the PPV of many biomarkers currently included in NBS, which has an aggregate PPV of 9 to 26%.^54,55^

### Lessons for variant reporting in gNBS

Certain P/LP variants were common among UKB and MGBB participants, the majority of whom had no clinical manifestations of the associated disorder. In particular, *RET* c.2410G>A (p.Val804Met) is a well-described low penetrance variant that accounted for approximately 15% of P/LP variants in both cohorts and is associated with an estimated lifetime risk of medullary thyroid cancer of approximately 4%.^56^ Although *RET* is commonly included in gNBS studies and is on the American College of Medical Genetics secondary findings list,^8,57^ excluding *RET* variants with known low penetrance may improve the PPV of gNBS and help avoid unnecessary interventions such as prophylactic thyroidectomy in children.

In contrast, some low penetrance or late-onset variants may prompt low-risk, confirmatory non-genetic testing, or lifesaving preventive care. A promoter variant in *OTC* (c.-106C>A), identified in one asymptomatic male in the MGBB, is associated with adult-onset symptoms.^58^ Because timely recognition of hyperammonemic crises prevents neurologic injury,^59^ it may be valuable to include all variants in *OTC* in screening, even if some individuals remain asymptomatic. Relatedly, a single variant in *F8*, c.6089G>A (p.Ser2030Asn) is associated with attenuated symptoms and accounted for 7-10% of P/LP variants in the UKB and MGBB. However, many such participants had suggestive clinical features of hemophilia.^60,61^ In individuals with this variant, clinical diagnosis can be corroborated by laboratory testing, enabling targeted management for individuals at risk of uncontrolled bleeding.^62^

More broadly, restricting the analysis of P/LP variants in X-linked conditions to chromosomal males could improve the PPV of gNBS. However, this approach risks missing females with attenuated symptoms who may benefit from anticipatory guidance and preventive care.^60,61^

### Limitations of biobank data

Although population biobanks provide large-scale genomic data, they underestimate the prevalence of P/LP variants because individuals with the most severe disease may be deceased or unable to participate in research.^24^ The UKB has been recognized as a source of low penetrance estimates, because in addition to participants often being healthy at enrollment due to volunteer bias, diagnostic codes captured in HESIN are limited to inpatient encounters.^13^

Additionally, the ICD codes used in this study correspond to the most severe and distinctive features of each condition, and therefore subtle manifestations, such as anxiety or depression in participants with *OTC* variants, may not be apparent.^63^ Estimated penetrance may also be falsely low when two assumed biallelic variants are later found to be in *cis*; however, most P/LP variants in this study occurred in X-linked or autosomal dominant genes, and those associated with autosomal recessive conditions were typically homozygous. Conversely, hospital-based biobanks may overestimate prevalence and severity because participants are often recruited while seeking medical care.^19^ The data provided in both biobanks may be incomplete if a participant sought care in healthcare systems outside of the accessed data.

### Other limitations

In addition to the limitations described above, this study has several other constraints. Both biobanks include mostly participants of European ancestry,^19,41^ which may result in underidentification of pathogenic variants among individuals of non-European descent, because variant interpretation resources are biased toward European populations.^64^ Misclassification of variants could lead to lower estimates of penetrance.^31^ For MGBB participants, sample swaps or sequencing artifacts could produce incorrect sequencing results,^19^ and symptom severity was assessed by only one medical geneticist. Only approximately 30% of eligible MGBB participants attended a clinical visit, limiting the ability to fully assess findings from these evaluations.

### Conclusions

As investigations of gNBS and the clinical use of genomic testing expand, clinicians need accurate estimates of the clinical impact of P/LP variants in asymptomatic individuals. Many genes associated with high-priority conditions for gNBS demonstrate a lifelong penetrance comparable to or exceeding the current PPV of NBS.^54^ Over time, biobank studies and gNBS programs, particularly those including participants from diverse populations, will be critical to refining estimates of variant penetrance and informing results reporting strategies for gNBS.

## Supporting information

eFigure 1

## Data Availability

All data produced in the present work are contained in the manuscript

## Acknowledgements

Artificial intelligence (ChatGPT) was used to draft initial lists of ICD codes (with research-in-the-loop review), draft code in R, and improve syntax and grammar.

## Author Contributions

Nina B. Gold had full access to all of the data in the study and takes responsibility for the integrity of the data and the accuracy of the data analysis.

*Conception and design: N.B.G., E.W.K., H.L.R., P.N., R.C.G., A.S., J.I.G*.

*Acquisition, analysis, or interpretation of data: N.B.G., H.Z., S.K., J.Y., E.P., M.S.S., L.O., H.S., E.M., A.C.F.L., S.L*.

*Drafting of the manuscript: N.B.G*.

*Critical revision of the manuscript for important intellectual content: N.B.G., H.Z., S.K., J.Y., E.P., M.S.S., L.O., H.S., E.M., A.C.F.L., E.W.K., H.L.R., P.N., R.C.G., S.L., A.S., J.I.G. Statistical analysis: N.B.G., H.Z., S.K., J.Y., M.S.S., S.L., A.S., J.I.G*.

*Obtaining funding: N.B.G., H.L.R., P.N., R.C.G*.

*Administrative, technical or material support: J.Y., E.P., L.O., H.S., E.M., A.C.F.L. Supervision: E.W.K., H.L.R., P.N., R.C.G*.

