## Supplement 1 for "Long-term Penetrance of Disease Variants in Genes Prioritized for Genomic Newborn Screening: Evidence from Adult Biobanks"

**TABLE OF CONTENTS**

| **eMethods** | **2** |
| --- | --- |
| **eTable 1.** List of genes and associated disorders queried in both the Mass General Brigham Biobank and U.K. Biobank. | **8** |
| **eTable 2**. Diagnostic ICD codes and ICD codes highly suggestive of the associated disease of U.K. and Mass General Brigham Biobank participants with pathogenic and likely pathogenic variants in an allelic state associated with disease. | **10** |
| **eTable 3.** Pathogenic and likely pathogenic variants identified in Mass General Brigham Biobank participants (n = 82) and their associated symptoms. | **16** |
| **eTable 4.** Pathogenic and likely pathogenic variants identified in U.K. Biobank participants (n = 665). | **34** |
| **eTable 5.** Participants in the U.K. Biobank with pathogenic and likely pathogenic variants in an allelic state that can cause disease with diagnostic ICD codes (n = 30). | **40** |
| **eTable 6.** Participants in the U.K. Biobank with pathogenic and likely pathogenic variants in an allelic state that can cause disease with ICD codes highly suggestive of the associated disease (n = 32). | **42** |
| **eTable 7.** Comparison of penetrance in Mass General Brigham Biobank participants, as measured by ICD codes and information in the notes of participants’ electronic medical records. | **44** |
| **eFigure 1.** REDCap data capture forms for the ten disorders associated with pathogenic and likely pathogenic variants identified in Mass General Brigham Biobank participants.***** |  |

### *Figure linked as a separate attachment.

### eMethods

**Assessment of P/LP variants in the U.K. Biobank and Mass General Brigham Biobank**

Exomes in both the UKB and MGBB were aligned to hg38. In both biobanks, all variants in the 54 genes of interest were identified, as described elsewhere.^1^ A medical geneticist (N.B.G.) then filtered these variants, retaining only those that had been previously classified as P/LP variants in ClinVar by at least one submitter, were not synonymous changes, had not been classified as benign by any submitter, or were putative loss-of-function variants with a high-confidence LOFTEE annotation and a minor allele frequency <1% in any gnomAD subpopulation.^2–4^ Variant classification was then performed in accordance with the criteria set by the guidelines by the American College of Medical Genetics (ACMG) and Association for Molecular Pathology,^5^ with modifications for individual conditions as recommended by the Clinical Genome Resource Expert Panels.^6^ Variants were subsequently reviewed by a molecular geneticist (H.Z.), who finalized the list of P/LP variants included in analyses (eTable 3 and eTable 6 in Supplement 1).

Participants harboring P/LP variants in an allelic state that can cause disease were then ascertained (i.e., one variant in genes associated with autosomal dominant or X-linked inheritance or two variants, not previously known to occur in *cis,**^4^* in genes associated with autosomal recessive inheritance). For recessive genes, variant phasing was assessed using gnomAD co-occurrence data and inspection of CRAM files (for MGBB participants) when variants were in close genomic proximity, enabling inference of *cis* versus *trans* configuration. Because X-linked conditions can manifest in both males and females, we identified participants of both chromosomal sexes with P/LP variants.^7,8^ Variants of uncertain significance were not sought or reported.

We described the demographic characteristics of participants in the UKB and MGBB who underwent exome sequencing, and among those harboring P/LP variants in an allelic state that can cause disease in the genes of interest. The chromosomal sex of UKB participants was inferred from X chromosome data: individuals with at least one heterozygous X chromosome genotype were classified as female and those with no heterozygous genotypes were classified as male. The sex of MGBB participants was derived from self-reported sex in the EMR. Analyses were completed using RStudio version 4.4.0 (2024-04-24) -- "Puppy Cup.”

**ICD-based phenotype ascertainment**

We defined “diagnostic ICD codes” as those that directly correspond to the genetic disorder of interest (e.g., an ICD code for hereditary factor VIII deficiency for participants with P/LP variants in *F8*). We defined ICD codes associated with highly suggestive symptoms as those that captured signs or symptoms that were strongly associated with the condition but not specific to it (e.g., an ICD code for hemarthrosis in participants with P/LP variants in *F8*). UKB participants with at least one of these codes were identified using Hospital Episode Statistics—Inpatient Diagnosis (HESIN) data. After identifying MGBB participants with P/LP variants, we identified the proportion who had the previously defined diagnostic ICD codes that directly correspond to the genetic disorder of interest and ICD codes for highly suggestive clinical features (eTable 2 in Supplement 1).

**EMR note abstraction in the Mass General Brigham Biobank**

Structured data collection forms for each genetic disease found in the MGBB were constructed using information from GeneReviews^9–19^ and primary literature on each condition (eFigure 1 in Supplement 1). We first determined whether participants with P/LP variants in an allelic state that can cause disease had a prior genetic diagnosis, defined as documentation in any EMR note of either the specific variant found in the MGBB data, or the name of the associated genetic condition. Using the structured data collection forms, a medical geneticist (N.B.G.) scored the severity of participant symptoms based on the detailed EMR review. Symptom severity was coded as: “no or limited symptoms,” for individuals with no apparent symptoms of disease or ubiquitous symptoms that were not necessarily due to the underlying monogenic variant; “moderate,” in individuals with at least one disease-related symptom that was suggestive of the underlying phenotype, or “marked,” for individuals with at least one pathognomonic finding related to the underlying monogenic disease. Symptom severity was encoded as “not enough information” when there were fewer than three encounters included in the EMR. Discrepancies in symptom severity coding were resolved through discussion between coders and occasionally consultation with content-area experts.

Differences in penetrance estimates between ICD code-based phenotyping and EMR review were assessed using McNemar’s test and Newcombe’s method for paired proportions. “ICD affected” was defined as participants who either had diagnostic ICD code(s) or ICD code(s) highly suggestive of the associated disease, and “ICD not affected” included those participants who did not have diagnostic code(s) or code(s) highly suggestive of the associated disease. “EMR affected” included participants who either had “Marked symptoms” or “Moderate symptoms” classifications in their pre-visit EMR review or a prior diagnosis recorded in the EMR notes, and “EMR not affected” included participants who had “No/limited symptoms” or “Not enough information in chart” classifications from their pre-visit EMR review and no prior diagnosis.

**MGBB participant recontact and clinical phenotyping**

Individuals who were living and had no genetic diagnosis recorded in their EMR were recontacted, first by an authorized letter from the MGBB and then by phone. Building on an existing IRB-approved protocol in the MGBB, if three consecutive phone calls were unanswered, the participant was considered to have passively opted out of participation. When reached by phone, the individual was given a broad explanation of the condition for which they were at risk (e.g., “a genetic condition that can cause the blood to not clot properly,” in regard to *F8*-related hemophilia). They were then offered to participate in a clinical visit with a medical geneticist. At the time of consent, they were informed that this clinical visit, as well as any associated laboratory or genetic testing performed during it, would be charged to their insurance. The participants were told that the data collected during this visit may be used retrospectively for research. All care conferred in the visit was clinical and for the benefit of the individual. At the targeted visits, targeted medical histories and laboratory testing was collected, and confirmatory clinical genetic testing was offered. Participants with suggestive clinical features were offered appropriate medical care.

### eTable 1. List of genes and associated disorders queried in both the Mass General Brigham Biobank and U.K. Biobank.

| **Gene** | **Disease** |
| --- | --- |
| Autosomal recessive | |
| *GBA* | *GBA*-related Gaucher disease |
| *CYP17A1* | *CYP17A1*-related 17-alpha-hydroxylase/17,20-lyase deficiency |
| *LIPA* | *LIPA*-related cholesteryl ester storage disease |
| *SLC37A4* | *SLC37A4*-related glycogen storage disease 1 |
| *ABCC8* | *ABCC8*-related hyperinsulinemic hypoglycemia |
| *G6PC* | *G6PC*-related glycogen storage disease type I |
| *CYP11B1* | *CYP11B1*-related congenital adrenal hyperplasia |
| *ARSB* | *ARSB*-related mucopolysaccharidosis type 6 |
| *GUSB* | *GUSB*-related mucopolysaccharidosis type 7 |
| *CPS1* | *CPS1*-related carbamoylphosphate synthetase I deficiency |
| *PLPBP* | *PLPBP*-related vitamin B6-dependent epilepsy |
| *ALDH7A1* | *ALDH7A1*-related pyridoxine-dependent epilepsy |
| *SLC26A3* | *SLC26A3*-related secretory diarrhea |
| *GATM* | *GATM*-related arginine:glycine amidinotransferase deficiency |
| *SLC7A7* | *SLC7A7*-related lysinuric protein intolerance |
| *NAGS* | *NAGS*-related N-acetylglutamate synthase deficiency |
| *AGL* | *AGL*-related glycogen storage disease 3 |
| *HSD3B2* | *HSD3B2*-related 3-beta-hydroxysteroid dehydrogenase 2 deficiency |
| *ALDOB* | *ALDOB*-related hereditary fructose intolerance |
| *PYGL* | *PYGL*-related glycogen storage disease 6 |
| *ATP7B* | *ATP7B*-related Wilson disease |
| *SLC9A3* | *SLC9A3*-related secretory diarrhea |
| *CTNS* | *CTNS*-related cystinosis |
| *FBP1* | *FBP1*-related fructose-1,6-bisphosphatase deficiency |
| *DGAT1* | *DGAT1*-related protein-losing enteropathy |
| *POMC* | *POMC*-related proopiomelanocortin deficiency |
| *MC2R* | *MC2R*-related glucocorticoid deficiency |
| *BCKDK* | *BCKDK*-related branched-chain keto acid dehydrogenase kinase deficiency |
| *MPI* | *MPI*-related congenital disorder of glycosylation (CDG Ib) |
| *PHKB* | *PHKB*-related glycogen storage disease 9 |
| *PHKG2* | *PHKG2*-related glycogen storage disease 9 |
| *SMPD1* | *SMPD1*-related acid sphingomyelinase deficiency |
| *GALNS* | *GALNS*-related mucopolysaccharidosis type 4 |
| *CYP11A1* | *CYP11A1*-related congenital adrenal insufficiency |
| *SLC25A15* | *SLC25A15*-related hyperornithinemia-hyperammonemia-homocitrullinemia syndrome |
| *GAMT* | *GAMT*-related guanidinoacetate methyltransferase deficiency |
| X-linked | |
| *F8* | *F8*-related hemophilia |
| *F9* | *F9*-related hemophilia |
| *IDS* | *IDS*-related mucopolysaccharidosis type 2 |
| *DMD* | *DMD*-related muscular dystrophy |
| *ATP7A* | *ATP7A*-related Menkes spectrum disorder |
| *PHKA2* | *PHKA2*-related glycogen storage disease 9 |
| *TAFAZZIN* | *TAFAZZIN*-related Barth syndrome |
| *PHKA1* | *PHKA1*-related glycogen storage disease, type IX |
| *OTC* | *OTC*-related ornithine transcarbamylase deficiency |
| *SLC6A8* | *SLC6A8*-related cerebral creatine deficiency |
| *GLA* | *GLA*-related Fabry disease |
| *BTK* | *BTK*-related agammaglobulinemia |
| Autosomal dominant | |
| *SLC2A1* | *SLC2A1*-related GLUT1 deficiency |
| *GLUD1* | *GLUD1*-related familial hyperinsulinism-hyperammonemia |
| *KCNJ11* | *KCNJ11*-related hyperinsulinemic hypoglycemia |
| *WT1* | *WT1*-related Wilms tumor |
| *RB1* | *RB1*-related retinoblastoma |
| *RET* | *RET*-related multiple endocrine neoplasia 2B |

### **eTable 2. Diagnostic ICD codes and ICD codes highly suggestive of the associated disease of U.K. and Mass General Brigham Biobank participants with pathogenic and likely pathogenic variants in an allelic state associated with disease.** (Abbreviations: MEN, multiple endocrine neoplasia; GI, gastrointestinal)

| **Gene** | **ICD Code** | **Description** |
| --- | --- | --- |
| *ATP7A* | E83.0 | Diagnosis: Disorders of copper metabolism |
| *ATP7A* | R62 | Lack of expected normal physiological development |
| *ATP7A* | R62.0 | Delayed milestone |
| *ATP7A* | R56 | Convulsions, not elsewhere classified |
| *ATP7A* | M62.81 | Other specified disorders of muscle (Shoulder region) |
| *BTK* | D80.1 | Diagnosis: Nonfamilial hypogammaglobulinaemia |
| *BTK* | D80.0 | Diagnosis: Hereditary hypogammaglobulinaemia |
| *BTK* | J18  J18.0  J18.1  J18.2  J18.8  J18.9 | Pneumonia, organism unspecified  Bronchopneumonia, unspecified  Lobar pneumonia, unspecified  Hypostatic pneumonia, unspecified  Other pneumonia, organism unspecified  Pneumonia, unspecified |
| *BTK* | J32  J32.0  J32.1  J32.2  J32.3  J32.4  J32.8  J32.9 | Chronic sinusitis  Chronic maxillary sinusitis  Chronic frontal sinusitis  Chronic ethmoidal sinusitis  Chronic sphenoidal sinusitis  Chronic pansinusitis  Other chronic sinusitis  Chronic sinusitis, unspecified |
| *BTK* | A41.9 | Septicaemia, unspecified |
| *DMD* | G71.0 | Diagnosis: Muscular dystrophy |
| *DMD* | M62.81 | Other specified disorders of muscle (Shoulder region) |
| *DMD* | M62.50 | Muscle wasting and atrophy, not elsewhere classified (Multiple sites) |
| *DMD* | Z99.3 | Dependence on wheelchair |
| *DMD* | J96.1 | Chronic respiratory failure |
| *DMD* | Z99.1 | Dependence on respirator |
| *DMD* | I42.0 | Dilated cardiomyopathy |
| *F8* | D66 | Diagnosis: Hereditary factor VIII deficiency |
| *F8* | R58 | Haemorrhage, not elsewhere classified |
| *F8* | N92.4 | Excessive bleeding in the premenopausal period |
| *F8* | R04 | Haemorrhage from respiratory passages |
| *F8* | M25.0  M25.00  M25.01  M25.02  M25.03  M25.04  M25.05  M25.06  M25.07  M25.08  M25.09 | Haemarthrosis  Haemarthrosis (Multiple sites)  Haemarthrosis (Shoulder region)  Haemarthrosis (Upper arm)  Haemarthrosis (Forearm)  Haemarthrosis (Hand)  Haemarthrosis (Pelvic region and thigh)  Haemarthrosis (Lower leg)  Haemarthrosis (Ankle and foot)  Haemarthrosis (Other)  Haemarthrosis (Site unspecified) |
| *F8* | Z14.01* | Asymptomatic hemophilia A carrier |
| *F9* | D67 | Diagnosis: Hereditary factor IX deficiency |
| *F9* | R58 | Haemorrhage, not elsewhere classified |
| *F9* | N924 | Excessive bleeding in the premenopausal period |
| *F9* | R04 | Haemorrhage from respiratory passages |
| *F9* | M25.0  M25.00  M25.01  M25.02  M25.03  M25.04  M25.05  M25.06  M25.07  M25.08  M25.09 | Haemarthrosis  Haemarthrosis (Multiple sites)  Haemarthrosis (Shoulder region)  Haemarthrosis (Upper arm)  Haemarthrosis (Forearm)  Haemarthrosis (Hand)  Haemarthrosis (Pelvic region and thigh)  Haemarthrosis (Lower leg)  Haemarthrosis (Ankle and foot)  Haemarthrosis (Other)  Haemarthrosis (Site unspecified) |
| *F9* | I61.9 | Intracerebral haemorrhage, unspecified |
| *F9* | I62.9 | Intracranial haemorrhage (nontraumatic), unspecified |
| *F9* | M79.8 | Other specified soft tissue disorders |
| *F9* | K92.0  K92.1  K92.2 | Haematemesis  Melaena  Gastro-intestinal haemorrhage, unspecified |
| *GLA* | E75.2 | Diagnosis: Other sphingolipidosis |
| *GLA* | G62 | G62 Other polyneuropathies |
| *GLA* | N18.1  N18.2  N18.3  N18.4  N18.5  N18.8  N18.9 | Chronic kidney disease, stage 1  Chronic kidney disease, stage 2  Chronic kidney disease, stage 3  Chronic kidney disease, stage 4  Chronic kidney disease, stage 5  Other chronic renal failure  Chronic renal failure, unspecified |
| *GLA* | I42 | Cardiomyopathy |
| *GLA* | I63.9 | Cerebral infarction, unspecified |
| *IDS* | E76.1 | Diagnosis: Mucopolysaccharidosis, type II |
| *IDS* | H90  H90.0  H90.1  H90.2  H90.3  H90.4  H90.5  H90.6  H90.7  H90.8 | Conductive and sensorineural hearing loss  Conductive hearing loss, bilateral  Conductive hearing loss, unilateral with unrestricted hearing on the contralateral side  Conductive hearing loss, unspecified  Sensorineural hearing loss, bilateral  Sensorineural hearing loss, unilateral with unrestricted hearing on the contralateral side  Sensorineural hearing loss, unspecified  Mixed conductive and sensorineural hearing loss, bilateral  Mixed conductive and sensorineural hearing loss, unilateral with unrestricted hearing on the contralateral side  Mixed conductive and sensorineural hearing loss, unspecified |
| *IDS* | G47.3 | Sleep apnoea |
| *IDS* | R62.0 | Delayed milestone |
| *IDS* | M24.5  M24.50  M24.51  M24.52  M24.53  M24.54  M24.55  M24.56  M24.57  M24.58  M24.59 | Contracture of joint  Contracture of joint (Multiple sites)  Contracture of joint (Shoulder region)  Contracture of joint (Upper arm)  Contracture of joint (Forearm)  Contracture of joint (Hand)  Contracture of joint (Pelvic region and thigh)  Contracture of joint (Lower leg)  Contracture of joint (Ankle and foot)  Contracture of joint (Other)  Contracture of joint (Site unspecified) |
| *IDS* | R16.0 | Hepatomegaly, not elsewhere classified |
| *IDS* | R16.1 | Splenomegaly, not elsewhere classified |
| *KCNJ11*** | E16.1  E16.2 | Other hypoglycemia  Hypoglycemia, unspecified |
| *KCNJ11* | E10.9  E10.65  E10.10  E10.11 | Type 1 diabetes mellitus without complications  Type 1 diabetes mellitus with hyperglycemia  Type 1 diabetes mellitus with ketoacidosis without coma  Type 1 diabetes mellitus with ketoacidosis with coma |
| *KCNJ11* | E13.9  E13.10  E13.11  E13.65  E13.8 | Other specified diabetes mellitus without complications  Other specified diabetes mellitus with ketoacidosis without coma  Other specified diabetes mellitus with ketoacidosis with coma  Other specified diabetes mellitus with hyperglycemia  Other specified diabetes mellitus with unspecified complications |
| *KCNJ11* | E11.65  E11.9 | Type 2 diabetes mellitus with hyperglycemia  Type 2 diabetes mellitus without complications |
| *KCNJ11* | R73.9 | Hyperglycemia, unspecified |
| *KCNJ11* | R73.03 | Prediabetes |
| *KCNJ11* | Z79.84 | Long term use of oral hypoglycemic drugs |
| *KCNJ11* | Z79.4 | Long term use of insulin |
| *OTC* | E72.4 | Diagnosis: Disorders of ornithine metabolism |
| *OTC* | G93.4 | Encephalopathy, unspecified |
| *OTC* | R41.8 | Other and unspecified symptoms and signs involving cognitive functions and awareness |
| *OTC* | G93.4 | Encephalopathy, unspecified |
| *PHKA1* | E74.0 | Diagnosis: Glycogen storage disease |
| *PHKA1* | R53 | Malaise and fatigue |
| *PHKA1* | M79.1  M79.10  M79.11  M79.12  M79.13  M79.14  M79.15  M79.16  M79.17  M79.18  M79.19 | Myalgia  Myalgia (Multiple sites)  Myalgia (Shoulder region)  Myalgia (Upper arm)  Myalgia (Forearm)  Myalgia (Hand)  Myalgia (Pelvic region and thigh)  Myalgia (Lower leg)  Myalgia (Ankle and foot)  Myalgia (Other)  Myalgia (Site unspecified) |
| *PHKA1* | M62.8 | Other specified disorders of muscle |
| *PHKA2* | E74.0 | Diagnosis: Glycogen storage disease |
| *PHKA2* | R16.0 | Hepatomegaly, not elsewhere classified |
| *PHKA2* | E16.2 | Hypoglycaemia, unspecified |
| *PHKA2* | R74.0 | Elevation of levels of transaminase and lactic acid dehydrogenase [LDH] |
| *SLC6A8* | E72.8 | Diagnosis: Other specified disorders of amino-acid metabolism |
| *SLC6A8* | F70  F70.0  F70.1  F70.8  F70.9 | Mild mental retardation  Mild mental retardation (With the statement of no, or minimal, impairment of behaviour)  Mild mental retardation (Significant impairment of behaviour requiring attention or treatment)  Mild mental retardation (Other impairments of behaviour)  Mild mental retardation (Without mention of impairment of behaviour) |
| *SLC6A8* | R56 | Convulsions, not elsewhere classified |
| *SLC6A8* | F84.0 | Childhood autism |
| *SLC6A8* | F80.9 | Developmental disorder of speech and language, unspecified |
| *TAFAZZIN* | E78 | Diagnosis: Disorders of lipoprotein metabolism and other lipidaemias |
| *TAFAZZIN* | I42.0 | Dilated cardiomyopathy |
| *TAFAZZIN* | D70 | Agranulocytosis |
| *TAFAZZIN* | R62 | Lack of expected normal physiological development |
| *TAFAZZIN* | R53 | Malaise and fatigue |
| *ALDOB* | E74.1 | Diagnosis: Disorders of fructose metabolism |
| *ALDOB* | E16.2 | Hypoglycaemia, unspecified |
| *ALDOB* | R11 | Nausea and vomiting |
| *ALDOB* | K76.9 | Liver disease, unspecified |
| *ALDOB* | R62 | Lack of expected normal physiological development |
| *ATP7B* | E83.0 | Diagnosis: Disorders of copper metabolism |
| *ATP7B* | K74.6 | Other and unspecified cirrhosis of liver |
| *ATP7B* | R17 | Unspecified jaundice |
| *ATP7B* | R25.1 | Tremor, unspecified |
| *ATP7B* | F06.8 | Other specified mental disorders due to brain damage and dysfunction and to physical disease |
| *MC2R* | E27.1 | Diagnosis: Primary adrenocortical insufficiency |
| *MC2R* | E16.2 | Hypoglycaemia, unspecified |
| *MC2R* | R62 | Lack of expected normal physiological development |
| *MC2R* | R53 | Malaise and fatigue |
| *RB1* | C69.2 | Diagnosis: Retina |
| *RB1* | Z85.8 | Personal history of malignant neoplasms of other organs and systems |
| *RB1* | H53.9 | Visual disturbance, unspecified |
| *RET* | E31 | Diagnosis: Polyglandular dysfunction |
| *RET* | C73 | Malignant neoplasm of thyroid gland |
| *RET* | Z85.8 | Personal history of malignant neoplasms of other organs and systems |
| *RET* | C74.1 | Medulla of adrenal gland |
| *RET* | D35.0 | Adrenal gland |
| *RET* | Q43.1 | Hirschsprung's disease |
| *SLC2A1* | E74.8 | Diagnosis: Other specified disorders of carbohydrate metabolism |
| *SLC2A1* | G40 | Epilepsy |
| *SLC2A1* | R56 | Convulsions, not elsewhere classified |
| *SLC2A1* | R26  R26.0  R26.1  R26.2  R26.3  R26.8 | Abnormalities of gait and mobility  Ataxic gait  Paralytic gait  Difficulty in walking, not elsewhere classified  Immobility  Other and unspecified abnormalities of gait and mobility |
| *SLC2A1* | F80.9 | Developmental disorder of speech and language, unspecified |
| *WT1* | N04.1 | Diagnosis: Focal and segmental glomerular lesions |
| *WT1* | C64 | Malignant neoplasm of kidney, except renal pelvis |
| *WT1* | N18.1  N18.2  N18.3  N18.4  N18.5  N18.8  N18.9 | Chronic kidney disease, stage 1  Chronic kidney disease, stage 2  Chronic kidney disease, stage 3  Chronic kidney disease, stage 4  Chronic kidney disease, stage 5  Other chronic renal failure  Chronic renal failure, unspecified |
| *WT1* | I10 | Essential (primary) hypertension |
| *WT1* | R60.9 | Oedema, unspecified |

*Code present only in MGBB data

**No diagnostic code for this disorder

### eTable 3. Pathogenic and likely pathogenic variants identified in Mass General Biobank participants (n = 82) and their associated symptoms. All variants were observed in a heterozygous state (or hemizygous for X-linked variants in males) unless otherwise noted. (Abbreviations: GERD, gastroesophageal reflux disease; GI, gastrointestinal; EMR, electronic medical record; ROM, range of motion; DDAVP, desmopressin; ITP, immune thrombocytopenic purpura; LV, left ventricle; MRI, magnetic resonance imaging; CAD, coronary artery disease; CABG, coronary artery bypass graft; CHF, congestive heart failure; TIA, transient ischemic attack; BPAD, bipolar affective disorder; PTSD, post-traumatic stress disorder; MS, multiple sclerosis; MEN2, multiple endocrine neoplasia type 2; MTC, medullary thyroid carcinoma; CEA, carcinoembryonic antigen; PTH, parathyroid hormone; ESRD, end-stage renal disease)

| **DNA change** | **Protein change** | **Chromosomal change** | **Gender** | **Pre-visit classificat-ion** | **Prior diagnosis in EMR** | **Clinical features abstracted from EMR** | **ICD Codes** | **Post-visit classificati-on** | **Additional clinical manifestations from visit** |
| --- | --- | --- | --- | --- | --- | --- | --- | --- | --- |
| ***ALDOB*-related hereditary fructose intolerance - Autosomal recessive (assumed compound heterozygous)** | | | | | | | | | |
| NM_000035.4:c.448G>C; NM_000035.4:c.524C>A | p.Ala150Pro; p.Ala175Asp | NC_000009.12:g.101427574C>G; NC_000009.12:g.101427498G>T | Female | Moderate symptoms | No | History of GERD, abdominal pain, nausea, and diarrhea following cholecystectomy. Coffee has been documented as a trigger of GI symptoms, but it is unclear if fructose is also a known trigger. |  | Marked symptoms | **Medical history:** Has always self-restricted sugar intake, even in childhood. Avoids all fruits or candies because she hates the taste. |
| ***DMD*-related muscular dystrophy - X-linked (hemizygous P/LP variants (males))** | | | | | | | | | |
| NM_004006.3:c.1812+1G>A | splice donor | NC_000023.11:g.32573529C>T | Male | Not enough information in chart | No | N/A |  | N/A |  |
| ***F8*-related hemophilia - X-linked (heterozygous P/LP variants (females) or hemizygous P/LP variants (males))** | | | | | | | | | |
| NM_000132.4:c.979C>G | p.Leu327Val | NC_000023.11:g.154969361G>C | Female | Marked symptoms | Yes | History of excessive bleeding after two procedures and frequent nosebleeds during childhood. | D66* | N/A |  |
| NM_000132.4:c.6104T>C | p.Val2035Ala | NC_000023.11:g.154902062A>G | Female | Marked symptoms | No | Developed a large hematoma of the breast two weeks after a breast reconstruction surgery and had a history of heavy menses. |  | Marked symptoms | **Medical history:** Lower extremity bruising.  **Family history:** Sister with a son with hemophilia A. |
| NM_000132.4:c.6506G>A | p.Arg2169His | NC_000023.11:g.154863151C>T | Male | Marked symptoms | Yes | Diagnosed with Hemophilia A at birth. History of severe hemophilic arthropathy with limited ROM in several joints. Had a surgery complicated by hematoma, despite factor VIII infusion. Very low baseline factor VIII levels. | D66*, R58, M25.00, M25.08 | N/A |  |
| NM_000132.4:c.6967C>T | p.Arg2323Cys | NC_000023.11:g.154837686G>A | Male | Marked symptoms | Yes | Diagnosed with Hemophilia A in childhood after a series of minor bleeds. Had excessive bleeding after dental extractions, sometimes requiring whole blood transfusions. Very low factor VIII levels. | D66* | N/A |  |
| NM_000132.4:c.575T>C | p.Ile192Thr | NC_000023.11:g.154992962A>G | Male | Marked symptoms | Yes | Diagnosed with Hemophilia A. Has experienced significant bleeding and bruising after procedures. Had a spontaneous hematoma in leg which resolved without factor VIII infusion. Moderately low factor VIII levels. |  | N/A |  |
| NM_000132.4:c.6506G>A | p.Arg2169His | NC_000023.11:g.154863151C>T | Female | Marked symptoms | Yes | Diagnosed with Hemophilia A in 20s after developing a large hematoma on forearm after minor trauma. Had frequent nosebleeds during childhood and had a history of easy bruising. Moderately low factor VIII levels. | D66*, Z14.01* | N/A |  |
| NM_000132.4:c.1649G>A | p.Arg550His | NC_000023.11:g.154957060C>T | Male | Marked symptoms | Yes | Diagnosed with Hemophilia A following excessive bleeding after dental extraction in childhood. History of frequent nosebleeds and bruising. History of excessive bleeding with surgeries. Hemarthrosis of knee. Moderately low baseline factor VIII levels. | D66*, M25.00 | N/A |  |
| NM_000132.4:c.5123G>A | p.Arg1708His | NC_000023.11:g.154928667C>T | Male | Marked symptoms | No | Diagnosed with Von Willebrand's disease in early adulthood due to increased clotting time of a cut on arm. VWB panel years later was normal. Had two episodes of significant hematomas. History of easy bruising on aspirin. Family history of excessive bruising. Slightly low factor VIII levels. | R58 | N/A |  |
| NM_000132.4:c.6089G>A | p.Ser2030Asn | NC_000023.11:g.154902077C>T | Female | Marked symptoms | Yes | Diagnosed with Von Willebrand's disease in childhood due to history of frequent nosebleeds, heavy menses, and easy bruising. Uses DDAVP with procedures, no bleeding complications. Also diagnosed with Hemophilia A after son's diagnosis. Moderately low baseline factor VIII levels. | D66* | N/A |  |
| NM_000132.4:c.2149C>T | p.Arg717Trp | NC_000023.11:g.154931641G>A | Female | Moderate symptoms | No | Mild hematoma following breast lumpectomy. Recurrent microscopic hematuria. |  | N/A |  |
| NM_000132.4:c.2149C>T | p.Arg717Trp | NC_000023.11:g.154931641G>A | Female | Moderate symptoms | No | Episode of GI bleeding. |  | N/A |  |
| NM_000132.4:c.6089G>A | p.Ser2030Asn | NC_000023.11:g.154902077C>T | Male | Moderate symptoms | No | Diagnosed with immune thrombocytopenic purpura. History of traumatic subarachnoid hemorrhage. History of frequent bruises and gingival bleeding. |  | Marked symptoms | **Medical history:** Subarachnoid hemorrhage was due to minimal trauma. Has frequent bruising and bleeding, including GI bleeding and worsening epistaxis.   **Family history:** Daughter with a history of bleeding.  **Laboratory results:** Elevated PTT and low Factor VIII activity. |
| NM_000132.4:c.6089G>A | p.Ser2030Asn | NC_000023.11:g.154902077C>T | Female | Moderate symptoms | No | History of frequent bruises and gingival bleeding. Father diagnosed with ITP. History of heavy menses. |  | Moderate symptoms | **Medical history:** History of gingival bleeding, easy bruising, and substantial bleeding following wisdom teeth removal.  **Family history:** Father with subarachnoid hemorrhage.  **Laboratory results:** Low factor VIII activity. |
| NM_000132.4:c.6089G>A | p.Ser2030Asn | NC_000023.11:g.154902077C>T | Female | Moderate symptoms | No | Excessive pain, bruising and swelling after ankle sprain, with ongoing swelling after seven weeks. |  | N/A |  |
| NM_000132.4:c.5399G>A | p.Arg1800His | NC_000023.11:g.154904998C>T | Female | No/limited symptoms | No | N/A |  | N/A |  |
| NM_000132.4:c.6089G>A | p.Ser2030Asn | NC_000023.11:g.154902077C>T | Female | No/limited symptoms | No | N/A (Known family history of hemophilia A) |  | N/A |  |
| NM_000132.4:c.6089G>A | p.Ser2030Asn | NC_000023.11:g.154902077C>T | Female | No/limited symptoms | No | N/A |  | No/limited symptoms | **Medical history:** No serious bleeding during previous procedures. History of heavy menses, which resolved. |
| NM_000132.4:c.6089G>A | p.Ser2030Asn | NC_000023.11:g.154902077C>T | Female | No/limited symptoms | No | N/A |  | N/A |  |
| NM_000132.4:c.6089G>A | p.Ser2030Asn | NC_000023.11:g.154902077C>T | Female | No/limited symptoms | No | N/A |  | N/A |  |
| ***F9*-related hemophilia - X-linked (heterozygous P/LP variants (females) or hemizygous P/LP variants (males))** | | | | | | | | | |
| NM_000133.4:c.148G>A | p.Gly50Ser | NC_000023.11:g.139537069G>A | Male | Marked symptoms | Yes | Diagnosed with Hemophilia B in childhood after bleeding from tonsillectomy. Has since taken prophylaxis before all procedures since diagnosis. Moderately low factor VIII levels. | D67* | N/A |  |
| NM_000133.4:c.572G>A | p.Arg191His | NC_000023.11:g.139551113G>A | Female | No/limited symptoms | No | N/A (Known family history of hemophilia B) |  | No/limited symptoms | **Medical history:** No bleeding episodes. History of one deep bruise took a prolonged time to resolve. History of heavy menses, but no history of anemia. **Family history:** Mother had a radical neck dissection and had a lot of uncontrolled bleeding. Sister has two sons with hemophilia. |
| NM_000133.4:c.572G>A | p.Arg191His | NC_000023.11:g.139551113G>A | Female | No/limited symptoms | No | N/A |  | N/A |  |
| ***GLA-*related Fabry disease - X-linked (heterozygous P/LP variants (females) or hemizygous P/LP variants (males))** | | | | | | | | | |
| NM_000169.3:c.132G>A | p.Trp44X | NC_000023.11:g.101407772C>T | Female | Marked symptoms | Yes | History of proteinuria. Diagnosed with Fabry disease after detection of corneal whorls. LV hypertrophy, angiokeratomas, white matter changes on MRI, intermittent joint pains, cold intolerance, hearing loss. Decreased GAL activity. | E75.21*, I42.5, I42.8, N18.31 | N/A |  |
| NM_000169.3:c.901C>G | p.Arg301Gly | NC_000023.11:g.101398468G>C | Male | Marked symptoms | Yes | Diagnosed with Fabry disease through genetic testing following an evaluation for hypertrophic cardiomyopathy. History of hypohidrosis, white matter changes on MRI. Chronic kidney disease. Early-onset hypertrophic cardiomyopathy, arrhythmias requiring pacemaker in 50s. Died from complications of heart failure. | E75.21*, I42.0, I42.1, I42.2, I42.5, I42.9, I42.8, N18.4, N18.5, N18.9, N18.30, N18.32, N18.3 | N/A |  |
| NM_000169.3:c.335G>A | p.Arg112His | NC_000023.11:g.101403845C>T | Male | Marked symptoms | Yes | Prior diagnosis through MGBB. History of CAD, underwent 4 CABG procedures, arrhythmias, LVH. History of hearing loss and tinnitus. Also history of pain in foot. Decreased GAL activity. | E75.21*, N18.30, N18.2, N18.3 | N/A |  |
| NM_000169.3:c.639+919G>A | NA | NC_000023.11:g.101399747C>T | Female | Marked symptoms | Yes | Prior diagnosis through MGBB. Acroparesthesia in hands and feet, which resolved after Rituximab infusions. History of vertigo and heat intolerance. Normal LV mass but low T1/ECV. | E75.21*, I42.2, I42.8, I42.9 | N/A |  |
| NM_000169.3:c.644A>G | p.Asn215Ser | NC_000023.11:g.101398942T>C | Female | Marked symptoms | Yes | History of acroparesthesias, GI symptoms since childhood. TIA symptoms with white matter changes on MRI. Mild hearing loss and tinnitus, hypohidrosis, small angiokeratomas. Poor tolerance to exercise and heat. | E75.21*, I42.5, I42.8 | N/A |  |
| NM_000169.3:c.427G>C | p.Ala143Pro | NC_000023.11:g.101401752C>G | Male | Marked symptoms | Yes | Diagnosed with Fabry disease in childhood, strong family history. History of severe GI symptoms, peripheral neuropathy, paresthesias, stroke and TIAs, severe hypertrophic cardiomyopathy, end stage renal disease resulting in kidney transplant, tinnitus and hearing problems. Died from complications of severe and progressive heart failure and kidney disease. | E75.21*, E75.22*, G62.9, I42.1, I42.2, I42.5, I42.9, I42.8, I42.0, I63.9, N18.30, N18.2, N18.32, N18.4, N18.6, N18.9, N18.5 | N/A |  |
| NM_000169.3:c.647A>G | p.Tyr216Cys | NC_000023.11:g.101398939T>C | Male | Marked symptoms | No | History of stroke and second episode of stroke-like symptoms. History of atrial fibrillation. Hearing loss. Labs show consistent mild proteinuria. |  | N/A |  |
| NM_000169.3:c.1184G>C | p.Gly395Ala | NC_000023.11:g.101397915C>G | Female | Marked symptoms | No | History of neuropathy in feet, exercise intolerance, vertigo. Cardiomyopathy leading to CHF. | I42.9, I42.8 | N/A |  |
| NM_000169.3:c.1184G>C | p.Gly395Ala | NC_000023.11:g.101397915C>G | Female | No/limited symptoms | Yes | Prior diagnosis through MGBB. No symptoms. | E75.21* | N/A |  |
| NM_000169.3:c.644A>G | p.Asn215Ser | NC_000023.11:g.101398942T>C | Female | No/limited symptoms | Yes | Prior diagnosis through MGBB. No symptoms. | E75.21*, I42.2 | N/A |  |
| NM_000169.3:c.335G>A | p.Arg112His | NC_000023.11:g.101403845C>T | Male | No/limited symptoms | No | N/A |  | N/A |  |
| NM_000169.3:c.1184G>C | p.Gly395Ala | NC_000023.11:g.101397915C>G | Female | No/limited symptoms | No | N/A | I42.9 | N/A |  |
| NM_000169.3:c.868A>C | p.Met290Leu | NC_000023.11:g.101398501T>G | Female | No/limited symptoms | Yes | Prior diagnosis through MGBB. Exhibits mild symptoms (vertigo, age-related hearing loss, tingling in feet), but was evaluated by a metabolic specialist and classified as not having specific symptoms related to Fabry disease. | E75.21*, G62.9, I42.2 | N/A |  |
| NM_000169.3:c.335G>A | p.Arg112His | NC_000023.11:g.101403845C>T | Female | No/limited symptoms | No | N/A |  | N/A |  |
| NM_000169.3:c.335G>A | p.Arg112His | NC_000023.11:g.101403845C>T | Female | No/limited symptoms | No | N/A |  | N/A |  |
| NM_000169.3:c.901C>G | p.Arg301Gly | NC_000023.11:g.101398468G>C | Female | No/limited symptoms | No | N/A |  | N/A |  |
| NM_000169.3:c.1184G>C | p.Gly395Ala | NC_000023.11:g.101397915C>G | Male | No/limited symptoms | No | Hearing loss related to environmental exposures. |  | N/A |  |
| ***KCNJ11*-related hyperinsulinemic hypoglycemia; *KCNJ11*-related permanent neonatal diabetes mellitus - Autosomal dominant (heterozygous P/LP variants)** | | | | | | | | | |
| NM_000525.4:c.101G>A | p.Arg34His | NC_000011.10:g.17387991C>T | Male | No/limited symptoms | No | N/A |  | N/A |  |
| NM_000525.4:c.101G>A | p.Arg34His | NC_000011.10:g.17387991C>T | Male | No/limited symptoms | No | N/A |  | No/limited symptoms | **New personal medical history:** No symptoms of hypoglycemia. Normal hemoglobin A1C.  **Family history:** One sibling died of SIDS. |
| NM_000525.4:c.616C>T | p.Arg206Cys | NC_000011.10:g.17387476G>A | Male | No/limited symptoms | No | N/A |  | N/A |  |
| NM_000525.4:c.901C>T | p.Arg301Cys | NC_000011.10:g.17387191G>A | Female | No/limited symptoms | No | N/A |  | N/A |  |
| ***OTC*-related ornithine transcarbamylase deficiency - X-linked (heterozygous P/LP variants (females) or hemizygous P/LP variants (males))** | | | | | | | | | |
| NM_000531.6:c.118C>T | p.Arg40Cys | NC_000023.11:g.38367331C>T | Female | Moderate symptoms | Yes | Prior diagnosis through MGBB. History of migraines that worsened during pregnancies. Diagnosed with depression and anxiety. Slightly elevated orotic acid levels. | E72.4* | N/A |  |
| NM_000531.6:c.817_819delGAG | p.Glu273del | NC_000023.11:g.38408972_38408974delGAG | Female | Moderate symptoms | No | Diagnosed with BPAD and PTSD, history of inpatient hospitalization due to psychosis. Has a shellfish allergy but no other known dietary restriction. |  | N/A |  |
| NM_000531.6:c.118C>T | p.Arg40Cys | NC_000023.11:g.38367331C>T | Female | Moderate symptoms | No | History of spontaneous abortion. Son with seizures in infancy. Personal history of anxiety and headaches. Allergy to shrimp/shellfish, no other known dietary restriction. |  | N/A |  |
| NM_000531.6:c.-106C>A | NA | NC_000023.11:g.38352591C>A | Male | No/limited symptoms | No | N/A |  | No/limited symptoms | **Medical history:** Tolerates high-protein meals well and does not avoid them.  **Family history:** Daughter has a learning disability. |
| ***PHKA1*-related glycogen storage disease, type IX - X-linked (heterozygous P/LP variants (females) or hemizygous P/LP variants (males))** | | | | | | | | | |
| NM_002637.4:c.2606+1G>A | splice donor | NC_000023.11:g.72609623C>T | Female | Marked symptoms | No | Chronic pain and numbness in extremities, particularly thigh, and lower back. Exercise intolerance and chronic fatigue. |  | N/A |  |
| NM_002637.4:c.2606+1G>A | splice donor | NC_000023.11:g.72609623C>T | Female | Marked symptoms | No | Proximal muscle weakness, pain in lower back exacerbated by activity. |  | N/A |  |
| NM_002637.4:c.2606+1G>A | splice donor | NC_000023.11:g.72609623C>T | Female | Marked symptoms | No | Neuropathy and muscle weakness in lower extremity since age 25. Difficulty ambulating and at risk for falls. History of muscle cramping and low exercise tolerance. |  | N/A |  |
| NM_002637.4:c.2606+1G>A | splice donor | NC_000023.11:g.72609623C>T | Female | Moderate symptoms | No | Some muscle pain reported in neck and shoulder. Abdominal muscle cramps and fatigue. | M62.838, M79.1 | Marked symptoms | **Medical history:** Muscle soreness for days following exercise or illness.  **Laboratory results:** Elevated creatine kinase. |
| NM_002637.4:c.2603_2604del | p.Ser868fs | NC_000023.11:g.72609625_72609626del | Female | Moderate symptoms | No | Exercise tolerance limited by chronic fatigue, diagnosed with MS. Episode of numbness in arm, leg. |  | N/A |  |
| NM_002637.4:c.2606+1G>A | splice donor | NC_000023.11:g.72609623C>T | Male | No/limited symptoms | No | N/A |  | N/A |  |
| NM_002637.4:c.2606+1G>A | splice donor | NC_000023.11:g.72609623C>T | Female | No/limited symptoms | No | N/A |  | No/limited symptoms | **Medical history:** Has some pain in the back and neck, but no severe pain in large muscle groups. No severe muscle pain during illnesses such as the flu. |
| NM_002637.4:c.2606+1G>A | splice donor | NC_000023.11:g.72609623C>T | Female | No/limited symptoms | No | N/A | R53.83 | N/A |  |
| NM_002637.4:c.2606+1G>A | splice donor | NC_000023.11:g.72609623C>T | Male | Not enough information in chart | No | N/A |  | N/A |  |
| ***RET*-related multiple endocrine neoplasia 2B - Autosomal dominant (heterozygous P/LP variants)** | | | | | | | | | |
| NM_020975.6:c.1858T>C | p.Cys620Arg | NC_000010.11:g.43113654T>C | Female | Marked symptoms | Yes | Family history of Hirschsprung's and MEN2. Diagnosed with MEN2 after discovering pheochromocytomas, leading to genetic testing. Had elevated plasma calcitonin and catecholamine levels, thyroid nodules that were discovered to be MTC during total thyroidectomy. | E31.22* C73, D35.00, Z85.850 | N/A |  |
| NM_020975.6:c.1852T>G | p.Cys618Gly | NC_000010.11:g.43113648T>G | Male | Marked symptoms | Yes | Family history of non-medullary thyroid cancer and breast cancer. Diagnosed with MEN2 after biopsy of neck mass showed MTC. Underwent total thyroidectomy and neck dissection to remove tumor. Elevated calcitonin and CEA. | E31.22*, C73, Z85.850 | N/A |  |
| NM_020975.6:c.1853G>A | p.Cys618Tyr | NC_000010.11:g.43113649G>A | Male | Marked symptoms | No | History of kidney stones. Mild hypercalcemia and elevated PTH consistent with primary hyperparathyroidism. Neck exploration revealed parathyroid adenoma, removed through partial parathyroidectomy. |  | Marked symptoms | No relevant new information collected |
| NM_020975.6:c.2410G>A | p.Val804Met | NC_000010.11:g.43119548G>A | Male | Marked symptoms | No | History of gastric neuroendocrine tumor and elevated PTH consistent with primary hyperthyroidism. |  | N/A |  |
| NM_020975.6:c.1858T>C | p.Cys620Arg | NC_000010.11:g.43113654T>C | Female | Marked symptoms | Yes | Family history of pheochromocytomas and MEN2/MTC. Diagnosed with MEN2A and Hirschsprung's in childhood. Had a total abdominal colectomy and ileostomy (later reversed) for enlarged bowel and constipation. History of kidney stones, hyperparathyroidism, elevated calcitonin. Had a total thyroidectomy and recurrent MTC, excised both times. | E31.22*, C73, Q43.1, Z85.850 | N/A |  |
| NM_020975.6:c.1901G>A | p.Cys634Tyr | NC_000010.11:g.43114501G>A | Male | Marked symptoms | Yes | Family history of MEN2 with MTC and pheochromocytoma. Personal medical history of MTC, pheochromocytomas, elevated calcitonin and CEA (now controlled), and pruritic cutaneous lichen amyloidosis. Had total thyroidectomy in childhood and partial adrenalectomy. | E31.22*, E31.21*, E31.23* , C73, C74.10, D35.00, D35.02, D35.01, Z85.850 | N/A |  |
| NM_020975.6:c.2671T>G | p.Ser891Ala | NC_000010.11:g.43120144T>G | Female | Moderate symptoms | No | No classic symptoms of MEN2. Strong family history of breast cancer (mother, father, and maternal aunt), as well as personal history of breast cancer. |  | N/A |  |
| NM_020975.6:c.2410G>A | p.Val804Met | NC_000010.11:g.43119548G>A | Male | Moderate symptoms | Yes | Did genetic testing for hereditary cancer risk due to personal history of prostate cancer and family members with breast/ovarian cancer. Found *RET* variant and children were also diagnosed. History of kidney stones and hyperparathyroidism. | E31.22* , C73, Z85.89 | N/A |  |
| NM_020975.6:c.2752A>G | p.Met918Val | NC_000010.11:g.43121967A>G | Female | Moderate symptoms | No | Family and personal history of papillary thyroid cancer, although no MTC. Had a total thyroidectomy after PTC was discovered. | C73, Z85.850 | Moderate symptoms | **Family history:** Two siblings with thyroid disease. None have had thyroid cancer or other cancers. |
| NM_020975.6:c.1900T>C | p.Cys634Arg | NC_000010.11:g.43114500T>C | Male | Moderate symptoms | Yes | Diagnosed after father underwent thyroidectomy for MTC. History of hyperparathyroidism and kidney stones, underwent parathyroidectomy to lower PTH levels. | E31.22*, Z85.850 | N/A |  |
| NM_020975.6:c.2410G>A | p.Val804Met | NC_000010.11:g.43119548G>A | Female | Moderate symptoms | No | Family and personal history of papillary thyroid cancer. Had a total thyroidectomy following PTC diagnosis. Also has extensive family history of cancer and underwent genetic testing for hereditary cancer, which was negative (but did not include RET). | C73, Z85.850 | N/A |  |
| NM_020975.6:c.2410G>A | p.Val804Met | NC_000010.11:g.43119548G>A | Female | Moderate symptoms | No | Did genetic testing due to significant history of cancer in her family and personal history. Personal history of recurrent breast cancer. Completed Invitae breast cancer panel (11 genes, but not RET), which was negative. No other symptoms of MEN2. | Z85.828 | No/limited symptoms | No relevant new information collected |
| NM_020975.6:c.2410G>A | p.Val804Met | NC_000010.11:g.43119548G>A | Female | Moderate symptoms | No | Family history of thyroid cancer (unknown type). Personal history of hyperparathyroidism and hypercalciuria, as well as breast cancer. No other symptoms of MEN2. |  | Moderate symptoms | **Family history:** Many siblings have had hyperthyroidism or hypothyroidism. |
| NM_020975.6:c.2410G>A | p.Val804Met | NC_000010.11:g.43119548G>A | Male | No/limited symptoms | No | N/A |  | N/A |  |
| NM_020975.6:c.2410G>A | p.Val804Met | NC_000010.11:g.43119548G>A | Female | No/limited symptoms | No | N/A |  | N/A |  |
| NM_020975.6:c.2410G>A | p.Val804Met | NC_000010.11:g.43119548G>A | Female | No/limited symptoms | No | N/A |  | N/A |  |
| NM_020975.6:c.2410G>A | p.Val804Met | NC_000010.11:g.43119548G>A | Female | No/limited symptoms | No | N/A |  | N/A |  |
| NM_020975.6:c.2410G>A | p.Val804Met | NC_000010.11:g.43119548G>A | Male | No/limited symptoms | No | N/A |  | N/A |  |
| NM_020975.6:c.1998G>C | p.Lys666Asn | NC_000010.11:g.43114598G>C | Male | No/limited symptoms | No | N/A |  | N/A |  |
| NM_020975.6:c.1998G>T | p.Lys666Asn | NC_000010.11:g.43114598G>T | Male | No/limited symptoms | No | N/A |  | N/A |  |
| NM_020975.6:c.2410G>A | p.Val804Met | NC_000010.11:g.43119548G>A | Female | No/limited symptoms | No | N/A |  | N/A |  |
| NM_020975.6:c.2410G>A | p.Val804Met | NC_000010.11:g.43119548G>A | Male | Not enough information in chart | No | N/A |  | N/A |  |
| NM_020975.6:c.2370G>C | p.Leu790Phe | NC_000010.11:g.43118458G>C | Female | Not enough information in chart | No | N/A |  | N/A |  |
| ***WT1*-related Wilms tumor - Autosomal dominant (heterozygous P/LP variants)** | | | | | | | | | |
| NM_024426.6:c.1447+4C>T | intron variant | NC_000011.10:g.32391968G>A | Female | Marked symptoms | No | Family history of two daughters with ESRD and diagnosis of Frasier syndrome (now accepted as one phenotypic representation of WT1 disorder). No personal diagnosis of Frasier syndrome, but has a history of ESRD and previous kidney transplant. History of persistent proteinuria, hypoalbuminemia, edema, hyperlipidemia, and diaphragmatic hernia. | I10, N18.32, N18.9, N18.3, N18.6, R60.9 | N/A |  |

* Diagnostic ICD code

### eTable 4. Pathogenic and likely pathogenic variants identified in U.K. Biobank participants. (Abbreviations: XL, X-linked; AR, autosomal recessive; AD, autosomal dominant)

| **Gene** | **Variant** | **Inheritance** | **N participants with variants in an allelic state that can cause disease** |
| --- | --- | --- | --- |
| *ATP7A* | chrX:77988478:C:G_C | XL | 1 |
| *ATP7A* | chrX:78012885:G:A_G | XL | 1 |
| *BTK* | chrX:101360567:C:T_C | XL | 1 |
| *BTK* | chrX:101375159:A:C_A | XL | 1 |
| *BTK* | chrX:101375202:C:T_C | XL | 1 |
| *DMD* | chrX:31134135:C:A_C | XL | 1 |
| *DMD* | chrX:31178798:G:C_G | XL | 1 |
| *DMD* | chrX:31204096:AC:A_AC | XL | 1 |
| *DMD* | chrX:31206663:G:A_G | XL | 1 |
| *DMD* | chrX:31348618:CG:C_CG | XL | 1 |
| *DMD* | chrX:31932083:G:GACTT_G | XL | 1 |
| *DMD* | chrX:32217065:T:C_T | XL | 1 |
| *DMD* | chrX:32365049:G:A_G | XL | 1 |
| *DMD* | chrX:32448611:CTT:C_CTT | XL | 1 |
| *DMD* | chrX:32468509:G:A_G | XL | 1 |
| *DMD* | chrX:32573744:C:T_C | XL | 1 |
| *DMD* | chrX:32697868:A:C_A | XL | 1 |
| *DMD* | chrX:32699117:G:A_G | XL | 1 |
| *DMD* | chrX:32809514:C:A_C | XL | 1 |
| *DMD* | chrX:32844794:G:A_G | XL | 1 |
| *DMD* | chrX:33211281:C:A_C | XL | 1 |
| *DMD* | chrX:33211281:C:T_C | XL | 1 |
| *DMD* | chrX:32348440:A:AC_A | XL | 2 |
| *DMD* | chrX:32472162:A:G_A | XL | 2 |
| *DMD* | chrX:32809559:G:A_G | XL | 2 |
| *DMD* | chrX:32844848:CT:C_CT | XL | 2 |
| *DMD* | chrX:32573529:C:T_C | XL | 49 |
| *F8* | chrX:154837697:G:A_G | XL | 1 |
| *F8* | chrX:154863151:C:T_C | XL | 1 |
| *F8* | chrX:154863223:A:C_A | XL | 1 |
| *F8* | chrX:154902084:C:T_C | XL | 1 |
| *F8* | chrX:154902120:G:A_G | XL | 1 |
| *F8* | chrX:154903942:C:CT_C | XL | 1 |
| *F8* | chrX:154904056:CA:C_CA | XL | 1 |
| *F8* | chrX:154928924:A:AT_A | XL | 1 |
| *F8* | chrX:154929410:AT:A_AT | XL | 1 |
| *F8* | chrX:154930313:AC:A_AC | XL | 1 |
| *F8* | chrX:154930488:TC:T_TC | XL | 1 |
| *F8* | chrX:154930615:T:A_T | XL | 1 |
| *F8* | chrX:154930666:CAATT:C_CAATT | XL | 1 |
| *F8* | chrX:154931623:C:T_C | XL | 1 |
| *F8* | chrX:154931640:C:T_C | XL | 1 |
| *F8* | chrX:154957073:G:A_G | XL | 1 |
| *F8* | chrX:154966077:G:A_G | XL | 1 |
| *F8* | chrX:154966425:C:T_C | XL | 1 |
| *F8* | chrX:154984719:G:A_G | XL | 1 |
| *F8* | chrX:154984733:C:T_C | XL | 1 |
| *F8* | chrX:154992962:A:G_A | XL | 1 |
| *F8* | chrX:154992996:C:T_C | XL | 1 |
| *F8* | chrX:154999531:TACAA:T_TACAA | XL | 1 |
| *F8* | chrX:154863125:G:A_G | XL | 2 |
| *F8* | chrX:154929919:C:CT_C | XL | 2 |
| *F8* | chrX:154961120:C:T_C | XL | 2 |
| *F8* | chrX:154966677:T:G_T | XL | 2 |
| *F8* | chrX:154837721:G:T_G | XL | 3 |
| *F8* | chrX:154861755:A:G_A | XL | 3 |
| *F8* | chrX:154928667:C:T_C | XL | 3 |
| *F8* | chrX:154931575:C:T_C | XL | 3 |
| *F8* | chrX:154947712:G:A_G | XL | 3 |
| *F8* | chrX:154984797:CT:C_CT | XL | 3 |
| *F8* | chrX:154863110:T:C_T | XL | 4 |
| *F8* | chrX:154953991:G:C_G | XL | 4 |
| *F8* | chrX:154863124:C:T_C | XL | 6 |
| *F8* | chrX:154953961:G:A_G | XL | 6 |
| *F8* | chrX:154969361:G:C_G | XL | 6 |
| *F8* | chrX:154957088:T:A_T | XL | 7 |
| *F8* | chrX:154903950:C:T_C | XL | 8 |
| *F8* | chrX:154931641:G:A_G | XL | 8 |
| *F8* | chrX:154902077:C:T_C | XL | 49 |
| *F9* | chrX:139537048:C:T_C | XL | 1 |
| *F9* | chrX:139537172:CAGTG:C_CAGTG | XL | 1 |
| *F9* | chrX:139541149:T:A_T | XL | 1 |
| *F9* | chrX:139548419:G:GA_G | XL | 1 |
| *F9* | chrX:139548420:ATAACAAGGTGG:A_ATAACAAGGTGG | XL | 1 |
| *F9* | chrX:139551060:A:G_A | XL | 1 |
| *F9* | chrX:139561710:C:T_C | XL | 1 |
| *F9* | chrX:139561835:C:CT_C | XL | 1 |
| *F9* | chrX:139537145:G:A_G | XL | 2 |
| *F9* | chrX:139561566:G:A_G | XL | 2 |
| *F9* | chrX:139561821:G:A_G | XL | 2 |
| *F9* | chrX:139537069:G:A_G | XL | 3 |
| *F9* | chrX:139541114:G:A_G | XL | 3 |
| *F9* | chrX:139561709:A:G_A | XL | 3 |
| *F9* | chrX:139537084:T:A_T | XL | 5 |
| *GLA* | chrX:101398467:C:T_C | XL | 1 |
| *GLA* | chrX:101398866:CTT:C_CTT | XL | 1 |
| *GLA* | chrX:101398907:G:A_G | XL | 1 |
| *GLA* | chrX:101398033:G:A_G | XL | 2 |
| *GLA* | chrX:101398891:A:G_A | XL | 3 |
| *GLA* | chrX:101398012:G:A_G | XL | 7 |
| *GLA* | chrX:101403845:C:T_C | XL | 10 |
| *GLA* | chrX:101398942:T:C_T | XL | 36 |
| *IDS* | chrX:149498301:G:A_G | XL | 1 |
| *OTC* | chrX:38403660:G:A_G | XL | 1 |
| *OTC* | chrX:38408987:C:T_C | XL | 1 |
| *OTC* | chrX:38403695:G:A_G | XL | 3 |
| *OTC* | chrX:38367331:C:T_C | XL | 8 |
| *OTC* | chrX:38403699:G:A_G | XL | 8 |
| *PHKA1* | chrX:72593131:A:C_A | XL | 1 |
| *PHKA1* | chrX:72593178:G:A_G | XL | 1 |
| *PHKA1* | chrX:72602010:GAC:G_GAC | XL | 1 |
| *PHKA1* | chrX:72602201:GT:G_GT | XL | 1 |
| *PHKA1* | chrX:72605337:G:A_G | XL | 1 |
| *PHKA1* | chrX:72605370:G:A_G | XL | 1 |
| *PHKA1* | chrX:72605389:T:TGTATGCATATG_T | XL | 1 |
| *PHKA1* | chrX:72611181:TC:T_TC | XL | 1 |
| *PHKA1* | chrX:72618850:C:T_C | XL | 1 |
| *PHKA1* | chrX:72620771:G:T_G | XL | 1 |
| *PHKA1* | chrX:72620775:G:GT_G | XL | 1 |
| *PHKA1* | chrX:72623107:A:G_A | XL | 1 |
| *PHKA1* | chrX:72644401:G:A_G | XL | 1 |
| *PHKA1* | chrX:72650389:C:A_C | XL | 1 |
| *PHKA1* | chrX:72650436:CA:C_CA | XL | 1 |
| *PHKA1* | chrX:72656194:TTTCAAATAGC:T_TTTCAAATAGC | XL | 1 |
| *PHKA1* | chrX:72657602:T:A_T | XL | 1 |
| *PHKA1* | chrX:72666266:G:GA_G | XL | 1 |
| *PHKA1* | chrX:72712805:T:A_T | XL | 1 |
| *PHKA1* | chrX:72712893:ATCTT:A_ATCTT | XL | 1 |
| *PHKA1* | chrX:72582562:C:A_C | XL | 2 |
| *PHKA1* | chrX:72584262:GAGGA:G_GAGGA | XL | 2 |
| *PHKA1* | chrX:72605280:G:A_G | XL | 2 |
| *PHKA1* | chrX:72623253:C:CT_C | XL | 2 |
| *PHKA1* | chrX:72644425:CA:C_CA | XL | 2 |
| *PHKA1* | chrX:72650401:ACATC:A_ACATC | XL | 2 |
| *PHKA1* | chrX:72650407:GGC:G_GGC | XL | 2 |
| *PHKA1* | chrX:72650460:TA:T_TA | XL | 2 |
| *PHKA1* | chrX:72712811:C:CA_C | XL | 2 |
| *PHKA1* | chrX:72611085:C:T_C | XL | 3 |
| *PHKA1* | chrX:72620832:AG:A_AG | XL | 3 |
| *PHKA1* | chrX:72635153:AC:A_AC | XL | 3 |
| *PHKA1* | chrX:72657614:G:A_G | XL | 3 |
| *PHKA1* | chrX:72609643:G:A_G | XL | 4 |
| *PHKA1* | chrX:72602196:G:A_G | XL | 5 |
| *PHKA1* | chrX:72605331:G:A_G | XL | 7 |
| *PHKA1* | chrX:72611075:G:A_G | XL | 7 |
| *PHKA2* | chrX:18897179:C:T_C | XL | 1 |
| *PHKA2* | chrX:18906802:T:TG_T | XL | 1 |
| *PHKA2* | chrX:18907017:C:T_C | XL | 1 |
| *PHKA2* | chrX:18908028:CAG:C_CAG | XL | 1 |
| *PHKA2* | chrX:18924436:G:T_G | XL | 1 |
| *PHKA2* | chrX:18925739:G:A_G | XL | 1 |
| *PHKA2* | chrX:18929276:AG:A_AG | XL | 1 |
| *PHKA2* | chrX:18931665:G:T_G | XL | 1 |
| *PHKA2* | chrX:18938673:C:T_C | XL | 1 |
| *SLC6A8* | chrX:153693312:TG:T_TG | XL | 1 |
| *SLC6A8* | chrX:153694448:T:A_T | XL | 1 |
| *SLC6A8* | chrX:153693312:T:TG_T | XL | 2 |
| *TAFAZZIN* | chrX:154420037:G:A_G | XL | 1 |
| *TAFAZZIN* | chrX:154420094:G:A_G | XL | 1 |
| *TAFAZZIN* | chrX:154420676:G:A_G | XL | 1 |
| *ALDOB* | chr9:101427574:C:G_C (homozygous) | AR | 12 |
| *ATP7B* | chr13:51941186:G:A_G (homozygous) | AR | 1 |
| *ATP7B* | chr13:51944145:G:T_G (homozygous) | AR | 2 |
| *ATP7B* | chr13:51944161:T:G_T (homozygous) | AR | 1 |
| *ATP7B* | chr13:51950132:C:T_C (homozygous) | AR | 6 |
| *ATP7B* | chr13:51975098:T:C_T (homozygous) | AR | 5 |
| *RB1* | chr13:48307278:A:G_A | AD | 1 |
| *RB1* | chr13:48347864:G:A_G | AD | 1 |
| *RB1* | chr13:48364893:G:A_G | AD | 1 |
| *RB1* | chr13:48379624:C:T_C | AD | 1 |
| *RB1* | chr13:48380197:CTT:C_CTT | AD | 1 |
| *RB1* | chr13:48381414:C:T_C | AD | 1 |
| *RB1* | chr13:48453032:C:T_C | AD | 1 |
| *RB1* | chr13:48456218:TA:T_TA | AD | 1 |
| *RB1* | chr13:48465238:C:T_C | AD | 1 |
| *RET* | chr10:43102421:G:A_G | AD | 1 |
| *RET* | chr10:43113613:A:G_A | AD | 1 |
| *RET* | chr10:43114491:G:T_G | AD | 1 |
| *RET* | chr10:43114501:G:A_G | AD | 1 |
| *RET* | chr10:43114737:G:A_G | AD | 1 |
| *RET* | chr10:43118392:G:C_G | AD | 1 |
| *RET* | chr10:43119548:G:C_G | AD | 1 |
| *RET* | chr10:43106497:G:A_G | AD | 2 |
| *RET* | chr10:43118392:G:T_G | AD | 2 |
| *RET* | chr10:43119602:GT:G_GT | AD | 2 |
| *RET* | chr10:43119606:GGC:G_GGC | AD | 2 |
| *RET* | chr10:43113622:G:T_G | AD | 3 |
| *RET* | chr10:43118458:G:C_G | AD | 3 |
| *RET* | chr10:43121967:A:G_A | AD | 3 |
| *RET* | chr10:43113622:G:A_G | AD | 6 |
| *RET* | chr10:43114596:A:G_A | AD | 8 |
| *RET* | chr10:43114598:G:C_G | AD | 8 |
| *RET* | chr10:43114598:G:T_G | AD | 8 |
| *RET* | chr10:43120144:T:G_T | AD | 9 |
| *RET* | chr10:43119548:G:T_G | AD | 14 |
| *RET* | chr10:43119548:G:A_G | AD | 111 |
| *SLC2A1* | chr1:42927685:G:A_G | AD | 1 |
| *SLC2A1* | chr1:42930766:G:A_G | AD | 1 |
| *SLC2A1* | chr1:42930865:G:A_G | AD | 1 |
| *WT1* | chr11:32396395:C:CAT_C | AD | 1 |
| *WT1* | chr11:32435186:G:A_G | AD | 1 |
| *WT1* | chr11:32435227:C:T_C | AD | 1 |
| *WT1* | chr11:32435179:C:CGG_C | AD | 2 |
| *WT1* | chr11:32417576:C:T_C | AD | 6 |

### eTable 5. Participants in the U.K. Biobank with pathogenic and likely pathogenic variants in an allelic state that can cause disease with diagnostic ICD codes (n = 30). (Abbreviations: XL, X-linked; AR, autosomal recessive; AD, autosomal dominant)

| **Gene** | **Variant** | **Mechanism of inheritance** | **Chromo-**  **somal sex** | **Other ICD codes highly suggestive of the associated disease** |
| --- | --- | --- | --- | --- |
| *DMD* | chrX:31348618:CG:C_CG | XL | Female |  |
| *DMD* | chrX:32573744:C:T_C | XL | Female |  |
| *F8* | chrX:154863151:C:T_C | XL | Male |  |
| *F8* | chrX:154929410:AT:A_AT | XL | Male |  |
| *F8* | chrX:154930488:TC:T_TC | XL | Male |  |
| *F8* | chrX:154931623:C:T_C | XL | Male | M25.07 |
| *F8* | chrX:154957073:G:A_G | XL | Female |  |
| *F8* | chrX:154966425:C:T_C | XL | Female |  |
| *F8* | chrX:154961120:C:T_C | XL | Male |  |
| *F8* | chrX:154953991:G:C_G | XL | Female |  |
| *F8* | chrX:154953961:G:A_G | XL | Female |  |
| *F8* | chrX:154953961:G:A_G | XL | Male |  |
| *F8* | chrX:154969361:G:C_G | XL | Male | R04.0 |
| *F8* | chrX:154957088:T:A_T | XL | Male |  |
| *F8* | chrX:154903950:C:T_C | XL | Male |  |
| *F9* | chrX:139561710:C:T_C | XL | Female |  |
| *F9* | chrX:139561821:G:A_G | XL | Female |  |
| *F9* | chrX:139561821:G:A_G | XL | Female |  |
| *GLA* | chrX:101398467:C:T_C | XL | Male | I42.1;I42.2;I42.9 |
| *GLA* | chrX:101398866:CTT:C_CTT | XL | Female |  |
| *GLA* | chrX:101398907:G:A_G | XL | Female | I42.2;I42.8;N18.5;N18.9 |
| *GLA* | chrX:101398891:A:G_A | XL | Male | I42.9 |
| *GLA* | chrX:101398891:A:G_A | XL | Male | N18.3;N18.9 |
| *GLA* | chrX:101398942:T:C_T | XL | Female |  |
| *GLA* | chrX:101398942:T:C_T | XL | Male | I42.1;I42.8 |
| *GLA* | chrX:101398942:T:C_T | XL | Male | I42.1;I42.2;I42.9;N18.3 |
| *OTC* | chrX:38403699:G:A_G | XL | Female |  |
| *ATP7B* | chr13:51944145:G:T_G | AR (homozygous) | Female | K74.6 |
| *RB1* | chr13:48381414:C:T_C | XL | Female | Z85.8 |
| *RB1* | chr13:48453032:C:T_C | XL | Female | Z85.8 |

### eTable 6. Participants in the U.K. Biobank with pathogenic and likely pathogenic variants in an allelic state that can cause disease with ICD codes highly suggestive of the associated disease (n = 32).

| **Gene** | **Variant** | **Mechanism of inheritance** | **Chromo-**  **somal sex** | **Diagnostic ICD code** | **Other ICD codes highly suggestive of the associated disease** |
| --- | --- | --- | --- | --- | --- |
| *DMD* | chrX:32573529:C:T_C | XL | Female | FALSE | Z991 |
| *DMD* | chrX:32573529:C:T_C | XL | Female | FALSE | I420 |
| *DMD* | chrX:32573529:C:T_C | XL | Male | FALSE | I420 |
| *F8* | chrX:154931623:C:T_C | XL | Male | TRUE | M2507 |
| *F8* | chrX:154837721:G:T_G | XL | Female | FALSE | R040 |
| *F8* | chrX:154969361:G:C_G | XL | Male | TRUE | R040 |
| *F8* | chrX:154969361:G:C_G | XL | Male | FALSE | R040 |
| *F8* | chrX:154902077:C:T_C | XL | Male | FALSE | R040 |
| *F9* | chrX:139551060:A:G_A | XL | Female | FALSE | K922 |
| *F9* | chrX:139541114:G:A_G | XL | Female | FALSE | K920 |
| *GLA* | chrX:101398467:C:T_C | XL | Male | TRUE | I421;I422;I429 |
| *GLA* | chrX:101398907:G:A_G | XL | Female | TRUE | I422;I428;N185;N189 |
| *GLA* | chrX:101398891:A:G_A | XL | Male | TRUE | I429 |
| *GLA* | chrX:101398891:A:G_A | XL | Male | TRUE | N183;N189 |
| *GLA* | chrX:101398942:T:C_T | XL | Male | TRUE | I421;I428 |
| *GLA* | chrX:101398942:T:C_T | XL | Male | FALSE | N183;N184;N189 |
| *GLA* | chrX:101398942:T:C_T | XL | Male | TRUE | I421;I422;I429;N183 |
| *ATP7B* | chr13:51944145:G:T_G | AR (homozygous) | Female | TRUE | K746 |
| *RB1* | chr13:48381414:C:T_C | AD | Female | TRUE | Z858 |
| *RB1* | chr13:48453032:C:T_C | AD | Female | TRUE | Z858 |
| *RET* | chr10:43114501:G:A_G | AD | Male | FALSE | Z858 |
| *RET* | chr10:43113622:G:A_G | AD | Female | FALSE | Z858 |
| *RET* | chr10:43119548:G:T_G | AD | Male | FALSE | Z858 |
| *RET* | chr10:43119548:G:A_G | AD | Male | FALSE | Z858 |
| *RET* | chr10:43119548:G:A_G | AD | Female | FALSE | C73 |
| *RET* | chr10:43119548:G:A_G | AD | Male | FALSE | Z858 |
| *RET* | chr10:43119548:G:A_G | AD | Female | FALSE | Z858 |
| *SLC2A1* | chr1:42930766:G:A_G | AD | Male | FALSE | R268 |
| *WT1* | chr11:32396395:C:CAT_C | AD | Male | FALSE | I10 |
| *WT1* | chr11:32435186:G:A_G | AD | Female | FALSE | I10;N189 |
| *WT1* | chr11:32417576:C:T_C | AD | Female | FALSE | I10 |
| *WT1* | chr11:32417576:C:T_C | AD | Male | FALSE | I10 |

### eTable 7. Comparison of penetrance in Mass General Brigham Biobank participants, as measured by ICD codes and information in the notes of participants’ electronic medical records. (Abbreviation: EMR, Electronic medical record; ICD, International Classification of Diseases)

|  | **EMR affected** | **EMR not affected** |
| --- | --- | --- |
| **ICD affected** | 30 | 2 |
| **ICD not affected** | 19 | 31 |
