## Supplementary material for "Long-term Penetrance of Disease Variants in Genes Prioritized for Genomic Newborn Screening: Evidence from Adult Biobanks": eFigure 1

### ALDOB Chart Review

Record ID ______________________

Variant ______________________

Medical Record Number ______________________

MGB Biobank Number ______________________

#### Demographic Information

Sex
- ◯ Male
- ◯ Female

Age ______________________

Race
- ☐ American Indian or Alaskan Native
- ☐ Asian Indian
- ☐ Black or African American
- ☐ East Asian
- ☐ Native Hawaiian or Pacific Islander
- ☐ Other
- ☐ White

Ethnicity
- ◯ Hispanic
- ◯ Not Hispanic

Dead/Alive
- ◯ Dead
- ◯ Alive

Cause of Death ______________________

Death related to variant?
- ◯ Yes
- ◯ No

#### Diagnosis

Hereditary Fructose Intolerance Diagnosis?
- ◯ Yes
- ◯ No

Age at diagnosis ______________________

Had genetic testing?
- ◯ Yes
- ◯ No

Type of genetic testing ______________________

Problem list

__________________________________

#### Family History

Family history

__________________________________

| | Yes | No |
|---|---|---|
| Diagnosis of HFI | ◯ | ◯ |
| Hypoglycemia | ◯ | ◯ |
| Chronic growth issues | ◯ | ◯ |
| Liver disease | ◯ | ◯ |
| Renal disease | ◯ | ◯ |
| Frequent lethargy or seizures | ◯ | ◯ |
| Adverse reaction to sweets | ◯ | ◯ |

#### Specialists Seen

Specialists seen

- ☐ Gastroenterology
- ☐ Nephrology
- ☐ Opthalmology
- ☐ Endocrinology
- ☐ Neurology
- ☐ Other Specialists

Last Gastrology A/P

__________________________________

Last Nephrology A/P

__________________________________

Last Opthalmalogy A/P

__________________________________

Last Endocrinology A/P

__________________________________

Last Neurology A/P

__________________________________

Other Specialists seen

__________________________________

#### Presence of HFI Symptoms

| | Yes | No |
|---|---|---|
| Reaction (i.e. sweating, vomiting) to fructose containing substances | ◯ | ◯ |
| Self restriction of fruit or sugar intake | ◯ | ◯ |
| Hypoglycemia | ◯ | ◯ |
| Lactic acidemia | ◯ | ◯ |
| Hypophosphatemia | ◯ | ◯ |
| Hyperuricemia | ◯ | ◯ |
| Hypermagnesemia | ◯ | ◯ |
| Hyperalaninemia | ◯ | ◯ |
| Lethargy | ◯ | ◯ |
| Diaphoresis | ◯ | ◯ |
| Frequent nausea | ◯ | ◯ |
| Frequent vomiting | ◯ | ◯ |
| Abdominal pain/distention | ◯ | ◯ |
| Ascites | ◯ | ◯ |
| Hepatomegaly | ◯ | ◯ |
| Hepatic dysfunction/failure | ◯ | ◯ |
| Hepatic adenoma or fibrosis | ◯ | ◯ |
| Non-alcoholic fatty liver disease | ◯ | ◯ |
| Renal insufficiency/failure | ◯ | ◯ |
| High glomerular filtration | ◯ | ◯ |
| High blood pressure | ◯ | ◯ |
| Chronic growth restriction | ◯ | ◯ |
| Seizures/coma as infant | ◯ | ◯ |
| Severe reactions to rotavirus vaccine | ◯ | ◯ |
| Lenticular cataracts | ◯ | ◯ |

#### Labs/Imaging/Procedures

Metabolic Testing

- ☐ Plasma glucose
- ☐ Plasma lactate
- ☐ Plasma phosphate
- ☐ Plasma uric acid
- ☐ Plasma magnesium
- ☐ Plasma alanine
- ☐ Carbohydrate deficient transferrin
- ☐ Other metabolite

Glucose Results

________________________________________

Lactate Results

__________________________________

Phosphate Results

__________________________________

Uric acid Results

__________________________________

Magnesium Results

__________________________________

Alanine Results

__________________________________

Carbohydrate deficient transferrin

__________________________________

Other metabolic test Results

__________________________________

Liver function tests

- ☐ Serum liver enzymes
- ☐ Coagulation factors
- ☐ Albumin
- ☐ Bilirubin
- ☐ Other liver function test

Liver enzyme result

__________________________________

Coagulation factors

__________________________________

Albumin results

__________________________________

Bilirubin results

__________________________________

Other liver function test

__________________________________

| | |
|---|---|
| Renal function test | ☐ BUN<br>☐ Serum creatinine<br>☐ Cystatin C<br>☐ Electrolyte levels<br>☐ Other renal function test |
| BUN Result | __________________________ |
| Serum creatinine result | __________________________ |
| Cystatin C result | __________________________ |
| Electrolyte Result | __________________________ |
| Other renal test result | __________________________ |
| Imaging | ☐ Renal US<br>☐ Abdominal US<br>☐ Abdominal CT/MRI<br>☐ Other Imaging |
| Renal US Results | __________________________ |
| Abdominal US Results | __________________________ |
| Abdominal CT/MRI Results | __________________________ |
| Other Imaging Results | __________________________ |
| Procedures | ☐ Liver Biopsy<br>☐ Endoscopy/GI Procedure<br>☐ Other Procedure |
| Liver Biopsy Results | __________________________ |
| Endoscopy/GI Procedure Results | __________________________ |

Other Procedure Results

__________________________________________

Other Comments

__________________________________________

Eligible for recontact?
- ◯ Yes
- ◯ No

Has symptoms of hereditary fructose intolerance?
- ◯ No/limited symptoms
- ◯ Moderate symptoms
- ◯ Marked symptoms
- ◯ Not enough information in chart

### DMD Chart Review

Record ID ______________________

Variant ______________________

Medical Record Number ______________________

MGB Biobank Number ______________________

#### Demographic Information

Sex
- ◯ Male
- ◯ Female

Age ______________________

Race
- ☐ American Indian or Alaskan Native
- ☐ Asian Indian
- ☐ Black or African American
- ☐ East Asian
- ☐ Native Hawaiian or Pacific Islander
- ☐ Other
- ☐ White

Ethnicity
- ◯ Hispanic
- ◯ Not Hispanic

Dead/Alive
- ◯ Dead
- ◯ Alive

Cause of Death ______________________

Death related to variant?
- ◯ Yes
- ◯ No

#### Diagnosis

Diagnosis
- ☐ DMD
- ☐ BMD
- ☐ DCM
- ☐ None

Age at diagnosis ______________________

Had genetic testing?
- ◯ Yes
- ◯ No

Type of genetic testing

__________________________________

Seeing genetics?
◯ Yes
◯ No

Problem list

__________________________________

#### Family History

Family history

__________________________________

#### Family History Suggestive of DMD Variant Conditions

| | Yes | No |
|---|---|---|
| Muscle weakness | ◯ | ◯ |
| Difficult walking, running, climbing stairs | ◯ | ◯ |
| Frequent falls | ◯ | ◯ |
| Waddling gait | ◯ | ◯ |
| Difficulty standing up | ◯ | ◯ |
| Early use of wheelchairs or mobility aids | ◯ | ◯ |
| Early onset cardiomyopathy | ◯ | ◯ |
| Early heart failure | ◯ | ◯ |
| Premature death | ◯ | ◯ |
| Recurrent muscle cramps | ◯ | ◯ |

#### Specialists Seen

Specialists seen
☐ Neurology
☐ Orthopedics
☐ Cardiology
☐ Pulmonology
☐ Physical Therapy
☐ Other specialists

Last Neurology A/P

__________________________________

Last Orthopedics A/P

__________________________________

Last Cardiology A/P

__________________________________

Last Pulmonology A/P

______________________________

Last Physical Therapy A/P

______________________________

Other Specialists seen

______________________________

#### Presence of DMD/BMD Symptoms

| | Yes | No |
|---|---|---|
| Low birth weight | ◯ | ◯ |
| Hyperglycemia | ◯ | ◯ |
| Developmental delay | ◯ | ◯ |
| Epilepsy | ◯ | ◯ |
| Muscle weakness | ◯ | ◯ |
| Hypotonia | ◯ | ◯ |
| Learning disabilities | ◯ | ◯ |
| Dystonia | ◯ | ◯ |
| Ataxia | ◯ | ◯ |
| Hyperactivity | ◯ | ◯ |
| Attention deficits | ◯ | ◯ |
| Glycosuria | ◯ | ◯ |
| Polyuria | ◯ | ◯ |
| Polydipsia | ◯ | ◯ |
| Failure to thrive in infancy | ◯ | ◯ |
| Dehydration | ◯ | ◯ |
| Ketoacidosis | ◯ | ◯ |
| Retinopathy | ◯ | ◯ |
| Nephropathy | ◯ | ◯ |
| Neuropathy | ◯ | ◯ |

If muscle weakness present, what was age of onset?

__________________________

#### Presence of DCM Symptoms

| | Yes | No |
|---|---|---|
| Hypoglycemia | ◯ | ◯ |
| Seizures | ◯ | ◯ |
| Hypotonia | ◯ | ◯ |

| | | |
|---|---|---|
| Poor feeding | ◯ | ◯ |
| Apnea | ◯ | ◯ |
| Lethargy | ◯ | ◯ |
| Weakness | ◯ | ◯ |
| Developmental delays | ◯ | ◯ |
| Poor weight gain | ◯ | ◯ |

#### Labs/Imaging/Procedures

Blood Testing

- ☐ Serum CK Levels
- ☐ Serum Myoglobin
- ☐ BNP Levels
- ☐ Other Test

Serum CK Results ______________________________

Serum Myoglobin Results ______________________________

BNP Results ______________________________

Other Blood Test Results ______________________________

Cardiac Tests/Procedures

- ☐ Echocardiogram
- ☐ Cardiac MRI
- ☐ Electrocardiogram
- ☐ Chest X-Ray
- ☐ Holter Monitoring
- ☐ Other Cardiac Test

Echocardiogram Results ______________________________

Cardiac MRI Results ______________________________

Electrocardiogram (ECG) Results ______________________________

Chest X-Ray Results ______________________________

Holter Monitoring Results ______________________________

| | |
|---|---|
| Other Cardiac Test Results | __________________________________ |
| Muscle Tests/Procedures | ☐ Muscle Biopsy<br>☐ Muscle MRI<br>☐ Electromyography<br>☐ Other Muscle Test |
| Muscle Biopsy Results | __________________________________ |
| Muscle MRI Results | __________________________________ |
| Electromyography Results | __________________________________ |
| Other Muscle Test Results | __________________________________ |
| Other Tests | ☐ Pulmonary Function Tests (PFTs)<br>☐ Bone Density Scan (DEXA)<br>☐ Other |
| Pulmonary Function Tests (PFTs) | __________________________________ |
| Bone Density Scan (DEXA) | __________________________________ |
| Other Test Results | __________________________________ |
| Other comments | __________________________________ |
| Eligible for recontact? | ◯ Yes<br>◯ No |
| Has symptoms of DMD/BMD/DCM? | ◯ No/limited symptoms<br>◯ Moderate symptoms<br>◯ Marked symptoms<br>◯ Not enough information in chart |

### F8 Chart Review

Record ID ______________________

Variant ______________________

Medical Record Number ______________________

MGB Biobank Number ______________________

#### Demographic Information

Sex
- ◯ Male
- ◯ Female

Age ______________________

Race
- ☐ American Indian or Alaskan Native
- ☐ Asian Indian
- ☐ Black or African American
- ☐ East Asian
- ☐ Native Hawaiian or Pacific Islander
- ☐ Other
- ☐ White

Ethnicity
- ◯ Hispanic
- ◯ Not Hispanic

Dead/Alive
- ◯ Dead
- ◯ Alive

Cause of Death ______________________

Death related to variant?
- ◯ Yes
- ◯ No

#### Diagnosis

Had genetic testing?
- ◯ Yes
- ◯ No

Hemophilia A diagnosis?
- ◯ Yes
- ◯ No

Age at genetic diagnosis ______________________

Type of genetic testing ______________________

Seeing genetics? ◯ Yes ◯ No

Problem list ______________________________

#### Family History

Family history ______________________________

#### Family History Suggestive of Hemophilia A

| | Yes | No |
|---|---|---|
| Known Hemophilia diagnosis | ◯ | ◯ |
| Prolonged bleeding | ◯ | ◯ |
| Hemoarthrosis | ◯ | ◯ |
| Postpartum hemorrhage | ◯ | ◯ |
| Intracranial bleeding | ◯ | ◯ |
| Excessive bruising | ◯ | ◯ |

#### Specialists Seen

Specialists seen
- ☐ Hematology
- ☐ Orthopedics
- ☐ Physical Therapy
- ☐ Other

Last Hematology A/P ______________________________

Last Ortho A/P ______________________________

Last Physical Therapy A/P ______________________________

Other Specialists seen ______________________________

#### Presence of Hemophilia A Symptoms

| | Yes | No |
|---|---|---|
| Prolonged bleeding after injuries, tooth extractions, surgery | ○ | ○ |
| Excessive pain and swelling from injuries, tooth extractions, surgery | ○ | ○ |
| Hemarthrosis | ○ | ○ |
| Hematomas | ○ | ○ |
| Intracranial bleeding | ○ | ○ |
| Neonatal cephalohematoma | ○ | ○ |
| GI bleeding | ○ | ○ |
| Hematuria | ○ | ○ |
| Heavy menstrual bleeding | ○ | ○ |
| Prolonged or recurrent nosebleeds | ○ | ○ |
| Excessive bruising | ○ | ○ |
| Firm subcutaneous hematomas | ○ | ○ |
| Spontaneous hemorrhage | ○ | ○ |
| Bleeding after circumcision | ○ | ○ |
| Bleeding from minor mouth injuries | ○ | ○ |
| Large "goose eggs" from minor head bumps | ○ | ○ |
| Joint pain | ○ | ○ |
| Joint swelling | ○ | ○ |
| Bleeding in muscles, kidneys, brain | ○ | ○ |
| Chronic joint disease | ○ | ○ |
| Postpartum hemorrhage | ○ | ○ |

#### Labs/Evaluations

Blood Testing

- ☐ CBC w/platelet count
- ☐ Activated Partial Thromboplastin Time (aPTT)
- ☐ Prothrombin Time (PT)
- ☐ Factor VIII Clotting Activity Test
- ☐ Von Willebrand Factor Level Test
- ☐ Alloimmune inhibitor screen
- ☐ Other blood test

CBC/Platelet Count Results

__________________________________

aPTT Results

__________________________________

| | |
|---|---|
| Prothrombin Results | __________________________________ |
| Factor VIII Clotting Activity Results | __________________________________ |
| Von Willebrand Factor Results | __________________________________ |
| Alloimmune inhibitor screen results | __________________________________ |
| Other blood test results | __________________________________ |
| Evaluations | ☐ Joint and muscle evaluation<br>☐ Other evaluation or test |
| Joint/muscle evaluation results | __________________________________ |
| Other Evaluation Results | __________________________________ |
| Other comments | __________________________________ |
| Eligible for recontact? | ◯ Yes<br>◯ No |
| Has symptoms of Hemophilia A? | ◯ No/limited symptoms<br>◯ Moderate symptoms<br>◯ Marked symptoms<br>◯ Not enough information in chart |

### F9 Chart Review

Record ID ______________________

Variant ______________________

Medical Record Number ______________________

MGB Biobank Number ______________________

#### Demographic Information

Sex
- ◯ Male
- ◯ Female

Age ______________________

Race
- ☐ American Indian or Alaskan Native
- ☐ Asian Indian
- ☐ Black or African American
- ☐ East Asian
- ☐ Native Hawaiian or Pacific Islander
- ☐ Other
- ☐ White

Ethnicity
- ◯ Hispanic
- ◯ Not Hispanic

Dead/Alive
- ◯ Dead
- ◯ Alive

Cause of Death ______________________

Death related to variant?
- ◯ Yes
- ◯ No

#### Diagnosis

Hemophilia B diagnosis?
- ◯ Yes
- ◯ No

Age at diagnosis ______________________

Had genetic testing?
- ◯ Yes
- ◯ No

Type of genetic testing ______________________

Problem list

__________________________________

#### Family History

Family history

__________________________________

#### Family History Suggestive of Hemophilia B

| | Yes | No |
|---|---|---|
| Known hemophilia diagnosis | ◯ | ◯ |
| Prolonged bleeding | ◯ | ◯ |
| Hemoarthrosis | ◯ | ◯ |
| Postpartum hemorrhage | ◯ | ◯ |
| Intracranial bleeding | ◯ | ◯ |
| Excessive bruising | ◯ | ◯ |

#### Specialists Seen

Specialists seen

- ☐ Hematology
- ☐ Orthopedics
- ☐ Physical Therapy
- ☐ Other

Last Hematology A/P

__________________________________

Last Ortho A/P

__________________________________

Last Physical Therapy A/P

__________________________________

Other Specialists seen

__________________________________

#### Presence of Hemophilia B Symptoms

| | Yes | No |
|---|---|---|
| Prolonged bleeding after injuries, tooth extractions, surgery | ◯ | ◯ |
| Excessive pain and swelling from injuries, tooth extractions, surgery | ◯ | ◯ |

| | | |
|---|---|---|
| Hemarthrosis | ○ | ○ |
| Deep-muscle hematomas | ○ | ○ |
| Intracranial bleeding | ○ | ○ |
| Neonatal cephalohematoma | ○ | ○ |
| GI bleeding | ○ | ○ |
| Hematuria | ○ | ○ |
| Heavy menstrual bleeding | ○ | ○ |
| Prolonged or recurrent nosebleeds | ○ | ○ |
| Excessive bruising | ○ | ○ |
| Firm subcutaneous hematomas | ○ | ○ |
| Spontaneous hemorrhage | ○ | ○ |
| Bleeding after circumcision | ○ | ○ |
| Bleeding from minor mouth injuries | ○ | ○ |
| Large "goose eggs" from minor head bumps | ○ | ○ |
| Joint pain | ○ | ○ |
| Joint swelling | ○ | ○ |
| Bleeding in muscles, kidneys, brain | ○ | ○ |
| Chronic joint disease | ○ | ○ |
| Postpartum hemorrhage | ○ | ○ |

#### Labs/Evaluations

Blood Testing
- ☐ CBC w/platelet count
- ☐ Activated Partial Thromboplastin Time (aPTT)
- ☐ Prothrombin Time (PT)
- ☐ Factor IX Clotting Activity Test
- ☐ Von Willebrand Factor Level Test
- ☐ Alloimmune inhibitor screen
- ☐ Other blood test

CBC/Platelet Count Results ________________________________

aPTT Results ________________________________

Prothrombin Results ________________________________

Factor IX Clotting Activity Results ________________________________

| | |
|---|---|
| Von Willebrand Factor Results | __________________________________ |
| Alloimmune inhibitor screen results | __________________________________ |
| Other blood test results | __________________________________ |
| Other comments | __________________________________ |
| Eligible for recontact? | ◯ Yes<br>◯ No |
| Has symptoms of Hemophilia B? | ◯ No/limited symptoms<br>◯ Moderate symptoms<br>◯ Marked symptoms<br>◯ Not enough information in chart |

### GLA Chart Review

Record ID ______________________________

Variant ______________________________

Medical Record Number ______________________________

MGB Biobank Number ______________________________

#### Demographic Information

Sex
- ◯ Male
- ◯ Female

Age ______________________________

Race
- ☐ American Indian or Alaskan Native
- ☐ Asian Indian
- ☐ Black or African American
- ☐ East Asian
- ☐ Native Hawaiian or Pacific Islander
- ☐ Other
- ☐ White

Ethnicity
- ◯ Hispanic
- ◯ Not Hispanic

Dead/Alive
- ◯ Dead
- ◯ Alive

Cause of Death ______________________________

Death related to variant?
- ◯ Yes
- ◯ No

#### Diagnosis

Fabry diagnosis?
- ◯ Yes
- ◯ No

Age at diagnosis ______________________________

Had genetic testing?
- ◯ Yes
- ◯ No

Type of genetic testing ______________________________

Seeing genetics?

◯ Yes
◯ No

Problem list

__________________________________________

#### Family History

Family history

__________________________________________

| | Yes | No |
|---|---|---|
| Fabry Diagnosis | ◯ | ◯ |
| Unexplained pain starting in childhood | ◯ | ◯ |
| Chronic Kidney Disease | ◯ | ◯ |
| Kidney Failure | ◯ | ◯ |
| Cardiomyopathy | ◯ | ◯ |
| Stroke or TIA | ◯ | ◯ |
| Neuropathic Pain | ◯ | ◯ |
| Abdominal pain without cause | ◯ | ◯ |
| Preeclampsia | ◯ | ◯ |
| Recurrent Miscarriages | ◯ | ◯ |
| Hearing Loss | ◯ | ◯ |

#### Specialists Seen

Specialists seen

☐ Ophthamology
☐ Nephrology
☐ Neurology
☐ Cardiology
☐ Dermatology
☐ Other Specialists

Last Ophtho A/P

__________________________________________

Last Nephro A/P

__________________________________________

Last Neuro A/P

__________________________________________

Last Cardio A/P

__________________________________________

Last Derm A/P

__________________________________

Other Specialists seen

__________________________________

#### Presence of Fabry Symptoms

| | Yes | No |
|---|---|---|
| Distal extremity pain/discomfort | ○ | ○ |
| Neuropathic pain | ○ | ○ |
| Exercise Intolerance | ○ | ○ |
| Cold intolerance | ○ | ○ |
| Heat intolerance | ○ | ○ |
| Hypohydrosis | ○ | ○ |
| Corneal problems | ○ | ○ |
| Proteinuria | ○ | ○ |
| Chronic kidney disease | ○ | ○ |
| s/p renal transplant | ○ | ○ |
| requiring dialysis | ○ | ○ |
| GI symptoms | ○ | ○ |
| Angiokeratomas | ○ | ○ |
| Cardiomyopathy | ○ | ○ |
| Aortic disease | ○ | ○ |
| Cardiac valve disease | ○ | ○ |
| Arrhythmia | ○ | ○ |
| Heart failure | ○ | ○ |
| Myocardial infarction | ○ | ○ |
| S/p stent placement or CABG | ○ | ○ |
| s/p ICD or pacemaker placement | ○ | ○ |
| s/p heart transplant or LVAD placement | ○ | ○ |
| Stroke | ○ | ○ |
| Vertigo | ○ | ○ |
| TIA | ○ | ○ |
| hearing loss | ○ | ○ |

#### Labs/Evaluations

Cardiac testing

- ☐ EKG
- ☐ Echocardiogram
- ☐ Stress test
- ☐ Cardiac MRI
- ☐ CTA chest
- ☐ Cardiac biopsy
- ☐ Troponin
- ☐ BNP
- ☐ Lipid panel
- ☐ Other

Results of last EKG

_______________________________

Results of last echocardiogram

_______________________________

Results of last stress test

_______________________________

Results of last cardiac MRI

_______________________________

Results of last CTA chest

_______________________________

Cardiac Biopsy

_______________________________

Troponin results

_______________________________

BNP levels (Max and most recent)

_______________________________

Last lipid panel results

_______________________________

Other cardiac results: what test and results

_______________________________

| | |
|---|---|
| Renal testing | ☐ UA<br>☐ Urine protein<br>☐ Urine microalbumin<br>☐ BMP<br>☐ Cystatin C<br>☐ Renal ultrasound<br>☐ Renal Biopsy<br>☐ Other |
| UA results | _______________________________ |
| Urine protein/creatinine ratio | _______________________________ |
| Urine microalbumin results | _______________________________ |
| Last serum Cr | _______________________________ |
| Max serum Cr | _______________________________ |
| Current GFR | _______________________________ |
| Cystatin C results | _______________________________ |
| Renal ultrasound results | _______________________________ |
| Renal biopsy results | _______________________________ |
| Other renal imaging: what test and results | _______________________________ |
| Neurological Testing | ☐ CT Brain<br>☐ MRI Brain<br>☐ MRA Head/Neck<br>☐ Carotid Ultrasound<br>☐ EMG/NCV<br>☐ Other Ultrasound (i.e. carpal tunnel)<br>☐ Other |

| | |
|---|---|
| CT head results | __________________________________ |
| MRI Brain results | __________________________________ |
| MRA head/neck results | __________________________________ |
| Carotid ultrasound results | __________________________________ |
| EMG/NCV results | __________________________________ |
| Other ultrasound: what US and results | __________________________________ |
| Other neuro tests: what test and results | __________________________________ |
| Other comments | __________________________________ |
| Eligible for recontact? | ◯ Yes<br>◯ No |
| Has symptoms of Fabry disease? | ◯ No/limited symptoms<br>◯ Moderate symptoms<br>◯ Marked symptoms<br>◯ Not enough information in chart |

### KCNJ11 Chart Review

| | |
|---|---|
| Record ID | __________________________________ |
| Variant | __________________________________ |
| Medical Record Number | __________________________________ |
| MGB Biobank Number | __________________________________ |

#### Demographic Information

| | |
|---|---|
| Sex | ◯ Male<br>◯ Female |
| Age | __________________________________ |
| Race | ☐ American Indian or Alaskan Native<br>☐ Asian Indian<br>☐ Black or African American<br>☐ East Asian<br>☐ Native Hawaiian or Pacific Islander<br>☐ Other<br>☐ White |
| Ethnicity | ◯ Hispanic<br>◯ Not Hispanic |
| Dead/Alive | ◯ Dead<br>◯ Alive |
| Cause of Death | __________________________________________ |
| Death related to variant? | ◯ Yes<br>◯ No |

#### Diagnosis

| | |
|---|---|
| Diagnosis | ☐ PNDM<br>☐ FHH<br>☐ None |
| Had genetic testing? | ◯ Yes<br>◯ No |
| Age at diagnosis | __________________________________ |
| Type of genetic testing | __________________________________ |

Seeing genetics? ◯ Yes ◯ No

Problem list ________________

#### Family History

Family history ________________

#### Family History Suggestive of KCNJ11 Variant Conditions

| | Yes | No |
|---|---|---|
| PNDM Diagnosis | ◯ | ◯ |
| FHH Diagnosis | ◯ | ◯ |
| Neonatal Diabetes | ◯ | ◯ |
| Early-Onset Diabetes | ◯ | ◯ |
| IUGR | ◯ | ◯ |
| Hypoglycemia | ◯ | ◯ |
| Insulin Resistance | ◯ | ◯ |
| Seizures in Infancy | ◯ | ◯ |
| Miscarriages/Stillbirths | ◯ | ◯ |
| Developmental Delays | ◯ | ◯ |

#### Specialists Seen

Specialists seen

- ☐ Endocrinology
- ☐ Nutrition
- ☐ Neurology
- ☐ Developmental Pediatrics
- ☐ Ophthamology
- ☐ Nephrology
- ☐ Other specialists

Last Endocrinology A/P ________________

Last Nutrition A/P ________________

Last Neurology A/P ________________

Last Dev Pediatrics A/P ________________

Last Ophthamology A/P

__________________________________________

Last Nephrology A/P

__________________________________________

Other Specialists seen

__________________________________________

#### Presence of PNDM Symptoms

| | Yes | No |
|---|---|---|
| Low birth weight | ◯ | ◯ |
| Hyperglycemia | ◯ | ◯ |
| IUGR | ◯ | ◯ |
| Developmental delay | ◯ | ◯ |
| Epilepsy | ◯ | ◯ |
| Muscle weakness | ◯ | ◯ |
| Hypotonia | ◯ | ◯ |
| Learning disabilities | ◯ | ◯ |
| Dystonia | ◯ | ◯ |
| Ataxia | ◯ | ◯ |
| Hyperactivity | ◯ | ◯ |
| Attention deficits | ◯ | ◯ |
| Glycosuria | ◯ | ◯ |
| Polyuria | ◯ | ◯ |
| Polydipsia | ◯ | ◯ |
| Failure to thrive in infancy | ◯ | ◯ |
| Dehydration | ◯ | ◯ |
| Ketoacidosis | ◯ | ◯ |
| Retinopathy | ◯ | ◯ |
| Nephropathy | ◯ | ◯ |
| Neuropathy | ◯ | ◯ |

#### Presence of CHI Symptoms

| | Yes | No |
|---|---|---|
| Hypoglycemia | ◯ | ◯ |
| Seizures | ◯ | ◯ |
| Hypotonia | ◯ | ◯ |
| Poor feeding | ◯ | ◯ |
| Apnea | ◯ | ◯ |

| | | |
|---|---|---|
| Lethargy | ○ | ○ |
| Weakness | ○ | ○ |
| Developmental delays | ○ | ○ |
| Poor weight gain | ○ | ○ |

#### Labs/Imaging/Procedures

Blood Testing

- ☐ Blood Glucose Test
- ☐ A1C
- ☐ Serum Insulin
- ☐ C-Peptide
- ☐ Oral Glucose Tolerance Test
- ☐ Urinalysis
- ☐ Autoantibody Testing
- ☐ Electrolyte Panel
- ☐ Lipid Panel
- ☐ Insulin Tolerance Test
- ☐ Glucagon Stimulation Test
- ☐ Other Test

Blood Glucose Test Results ______________________________

A1C Results ______________________________

Serum Insulin Results ______________________________

C-Peptide Results ______________________________

Oral Glucose Tolerance Test Results ______________________________

Urinalysis Results ______________________________

Autoantibody Test Results ______________________________

Electrolyte panel Results ______________________________

Lipid Panel Results ______________________________

Insulin Tolerance Test Results

__________________________________

Glucagon Stimulation Test Results

__________________________________

Other Blood Test Results

__________________________________

Procedures

- ☐ Neurodevelopmental Assessments
- ☐ Pancreatic Procedure
- ☐ Other Procedure

Neurodevelopmental Assessments Results

__________________________________

Pancreatic Procedure Results

__________________________________

Other Procedure Results

__________________________________

Other comments

__________________________________

Eligible for recontact?

- ◯ Yes
- ◯ No

Has symptoms of FHH or PNDM?

- ◯ No/limited symptoms
- ◯ Moderate symptoms
- ◯ Marked symptoms
- ◯ Not enough information in chart

### OTC Chart Review

Record ID ______________________

Variant ______________________

Medical Record Number ______________________

MGB Biobank Number ______________________

#### Demographic Information

Sex
- ◯ Male
- ◯ Female

Age ______________________

Race
- ☐ American Indian or Alaskan Native
- ☐ Asian Indian
- ☐ Black or African American
- ☐ East Asian
- ☐ Native Hawaiian or Pacific Islander
- ☐ Other
- ☐ White

Ethnicity
- ◯ Hispanic
- ◯ Not Hispanic

Dead/Alive
- ◯ Dead
- ◯ Alive

Cause of Death ______________________

Death related to variant?
- ◯ Yes
- ◯ No

#### Diagnosis

OTC deficiency diagnosis?
- ◯ Yes
- ◯ No

Age at diagnosis ______________________

Had genetic testing?
- ◯ Yes
- ◯ No

Type of genetic testing ______________________

Seeing genetics?

◯ Yes
◯ No

Problem list

____________________________________

#### Family History

Family history

____________________________________

| | Yes | No |
|---|---|---|
| Known OTC diagnosis | ◯ | ◯ |
| Urea cycle disorders | ◯ | ◯ |
| Unexplained newborn male death | ◯ | ◯ |
| Cerebral palsy NOS | ◯ | ◯ |
| Migraines/Headaches | ◯ | ◯ |
| Psychiatric conditions | ◯ | ◯ |
| Recurrent vomiting | ◯ | ◯ |
| Other decompensation during hospitalization/stress | ◯ | ◯ |

#### Pregnancy History

Ever pregnant?

◯ Yes
◯ No

Living children. List as (#M, #F)

____________________________________

#### Other pregnancy history suggestive of OTC deficiency

| | Yes | No |
|---|---|---|
| Spontaneous abortion | ◯ | ◯ |
| Had newborn male child who died | ◯ | ◯ |
| Had male child with sepsis in first week of life | ◯ | ◯ |
| Had male child with failure to thrive in first week of liife | ◯ | ◯ |
| Had male child with somnolence in first week of life | ◯ | ◯ |
| Had a male child with unexplained tachypnea in first week of life | ◯ | ◯ |

| | | |
|---|---|---|
| Elective abortion | ○ | ○ |

Other pregnancy history ______________________

#### Specialists Seen

Specialists seen
- ☐ Psychiatry
- ☐ Neurology
- ☐ OB/GYN
- ☐ GI
- ☐ Nutrition
- ☐ Other Specialists

Last Psychiatry A/P ______________________

Last Neurology A/P ______________________

Last OB/GYN A/P ______________________

Last GI A/P ______________________

Last Nutrition A/P ______________________

Other Specialists seen ______________________

#### Presence of OTC Deficiency Symptoms

| | Yes | No |
|---|---|---|
| Delirium | ○ | ○ |
| Ecephalopathy or Altered Mental Status | ○ | ○ |
| Psychosis or erratic behavior NOS | ○ | ○ |
| Anxiety/Depression | ○ | ○ |
| Recurrent vomiting | ○ | ○ |
| Headaches/Migraine headaches | ○ | ○ |
| Seizure | ○ | ○ |

| | | |
|---|---|---|
| Allergy to protein/protein intolerance | ○ | ○ |
| HCC | ○ | ○ |
| Reye-like syndrome | ○ | ○ |
| Executive function deficits | ○ | ○ |
| Mild cognitive impairment | ○ | ○ |

Self-restriction of protein / vegetarian diet / avoidance of milk, red meat, eggs, high-protein foods (list specific food restrictions) ________________________________

Headache description (From most recent PCP A/P Problem list) ________________________________

#### Presence of OTC deficiency symptoms during precipitating events or catabolic stress

| | Altered Mental Status | Vomiting | Headache |
|---|---|---|---|
| Pregnancy and/or Delivery | ☐ | ☐ | ☐ |
| Systemic corticosteroid administration | ☐ | ☐ | ☐ |
| Infection | ☐ | ☐ | ☐ |
| Trauma (e.g. motor vehicle collision) | ☐ | ☐ | ☐ |
| Cancer therapy | ☐ | ☐ | ☐ |
| Meat/protein ingestion | ☐ | ☐ | ☐ |

#### Labs/Evaluations

Neurologic testing
- ☐ EEG
- ☐ Head CT
- ☐ Head MRI
- ☐ Other neuropsych testing

Results of any abnormal EEG ________________________________________

Head CT Results ________________________________________

Head MRI Results ________________________________________

Other neuropsych testing and results ________________________________________

Other Imaging
- ☐ Abdominal US
- ☐ CT A/P
- ☐ Other Liver imaging

Results of last abdominal U/S

______________________________

Results of last CT A/P (Living findings and impression)

______________________________

Other liver imaging: what test and results

______________________________

Other labs

- ☐ Ammonia (serum)
- ☐ Blood gas
- ☐ LFTs
- ☐ PT/PTT
- ☐ Urine studies

Serum ammonia levels with dates

______________________________

ABG showing respiratory alkalosis?

- ◯ Yes
- ◯ No

Last AST/ALT

______________________________

Last PT/PTT

______________________________

Elevated urine orotic acid

______________________________

Other comments

______________________________

Eligible for recontact?

- ◯ Yes
- ◯ No

Has symptoms of OTC deficiency?

- ◯ No/limited symptoms
- ◯ Moderate symptoms
- ◯ Marked symptoms
- ◯ Not enough information in chart

### PHKA1 Chart Review

Record ID ________________

Variant ________________

Medical Record Number ________________

MGB Biobank Number ________________

#### Demographic Information

Sex
- ◯ Male
- ◯ Female

Age ________________

Race
- ☐ American Indian or Alaskan Native
- ☐ Asian Indian
- ☐ Black or African American
- ☐ East Asian
- ☐ Native Hawaiian or Pacific Islander
- ☐ Other
- ☐ White

Ethnicity
- ◯ Hispanic
- ◯ Not Hispanic

Dead/Alive
- ◯ Dead
- ◯ Alive

Cause of Death ________________

Death related to variant?
- ◯ Yes
- ◯ No

#### Diagnosis

Had genetic testing?
- ◯ Yes
- ◯ No

GSD diagnosis?
- ◯ Yes
- ◯ No

Age at genetic diagnosis ________________

Type of genetic testing ________________

Seeing genetics?

◯ Yes
◯ No

Problem list

__________________

#### Family History

Family history

__________________

#### Family History Suggestive of GSD

| | Yes | No |
|---|---|---|
| Known GSD diagnosis | ◯ | ◯ |
| Exercise intolerance | ◯ | ◯ |
| Elevated creatine kinase | ◯ | ◯ |
| Rhabdomyolysis | ◯ | ◯ |
| Abnormal response to anaesthesia | ◯ | ◯ |
| Statin-induced myopathy | ◯ | ◯ |

#### Specialists Seen

Specialists seen

☐ Metabolics
☐ Neurology
☐ Endocrinology
☐ Physical Therapy
☐ Orthopedics
☐ Other

Last Metabolics A/P

__________________

Last Neuro A/P

__________________

Last Endocrine A/P

__________________

Last PT A/P

__________________

Last Ortho A/P

__________________

Other Specialists Seen

______________________________________

#### Presence of GSD Symptoms

| | Yes | No |
|---|---|---|
| Muscle weakness | ◯ | ◯ |
| Weakness in pelvic girdle/lower limbs | ◯ | ◯ |
| Exercise intolerance | ◯ | ◯ |
| Muscular atrophy | ◯ | ◯ |
| Myalgia | ◯ | ◯ |
| Muscle stiffness | ◯ | ◯ |
| Muscle cramps | ◯ | ◯ |
| Fatigue | ◯ | ◯ |
| Muscle fiber necrosis | ◯ | ◯ |
| Elevated creatine kinase | ◯ | ◯ |
| Increased muscle glycogen | ◯ | ◯ |
| Myoglobinuria | ◯ | ◯ |
| Reduced muscle phosphorylase kinase | ◯ | ◯ |
| Rhabdomyolysis | ◯ | ◯ |
| Abnormal response to anaesthesia | ◯ | ◯ |
| Statin-induced myopathy | ◯ | ◯ |

#### Labs/Evaluations

Blood Testing

- ☐ Creatine Kinase (CK)†
- ☐ Blood Glucose
- ☐ Serum Lactate
- ☐ Serum Electrolytes
- ☐ Other blood test

Creatine Kinase (CK) Levels

______________________________________

Blood Glucose Levels

______________________________________

Serum Lactate Levels

______________________________________

Serum Electrolytes Results

______________________________________

Other blood test results

__________________________________

Procedures/Imaging

- ☐ Muscle Biopsy
- ☐ Glycogen Content Assay
- ☐ PhK Activity Assay
- ☐ Electromyography
- ☐ Muscle Ultrasound/MRI
- ☐ Exercise Testing
- ☐ Other Test

Muscle Biopsy Results

__________________________________

Glycogen Content Assay Results

__________________________________

PhK Activity Assay Results

__________________________________

Electromyography (EMG) Results

__________________________________

Muscle Ultrasound/MRI Results

__________________________________

Exercise Testing Results

__________________________________

Other Evaluation Results

__________________________________

Other comments

__________________________________

Eligible for recontact?

- ◯ Yes
- ◯ No

Has symptoms of Glycogen Storage Disease?

- ◯ No/limited symptoms
- ◯ Moderate symptoms
- ◯ Marked symptoms
- ◯ Not enough information in chart

### RET Chart Review

Record ID ______________________

Variant ______________________

Medical Record Number ______________________

MGB Biobank Number ______________________

#### Demographic Information

Sex
- ◯ Male
- ◯ Female

Age ______________________

Race
- ☐ American Indian or Alaskan Native
- ☐ Asian Indian
- ☐ Black or African American
- ☐ East Asian
- ☐ Native Hawaiian or Pacific Islander
- ☐ Other
- ☐ White

Ethnicity
- ◯ Hispanic
- ◯ Not Hispanic

Dead/Alive
- ◯ Dead
- ◯ Alive

Cause of Death ______________________

Death related to variant?
- ◯ Yes
- ◯ No

#### Diagnosis

Had genetic testing?
- ◯ Yes
- ◯ No

Diagnosis
- ☐ MEN2
- ☐ FMTC
- ☐ Hirschsprung Disease
- ☐ PTC
- ☐ None

Age at genetic diagnosis ______________________

Type of genetic testing

__________________________________

Seeing genetics?

◯ Yes
◯ No

Problem list

__________________________________

#### Family History

Family history

__________________________________

#### Family History Suggestive of RET Variant Conditions

| | Yes | No |
|---|---|---|
| Known MEN2 Diagnosis | ◯ | ◯ |
| Early tumor development | ◯ | ◯ |
| Medullary Thyroid Carcinoma | ◯ | ◯ |
| Pheochromocytoma | ◯ | ◯ |
| Hyperparathyroidism | ◯ | ◯ |
| Mucosal Neuroma | ◯ | ◯ |
| Marfanoid habitus | ◯ | ◯ |
| Hirschsprung Disease | ◯ | ◯ |
| Recurrent constipation | ◯ | ◯ |

Type of tumor/onset

__________________________________

#### Specialists Seen

Specialists seen

- ☐ Endocrinology
- ☐ Oncology
- ☐ Gastroenterology
- ☐ Ophthalmology
- ☐ Cardiology
- ☐ Dermatology
- ☐ Other specialists

Last Endocrinology A/P

__________________________________

Last Oncology A/P

__________________________________

Last Gastroenterology A/P

__________________________________

Last Ophthalmology A/P

__________________________________

Last Cardiology A/P

__________________________________

Last Dermatology A/P

__________________________________

Other Specialists seen

__________________________________

#### Presence of MEN2A Symptoms

| | Yes | No |
|---|---|---|
| Medullary thyroid carcinoma | ○ | ○ |
| Neck mass | ○ | ○ |
| Neck pain | ○ | ○ |
| Diarrhea | ○ | ○ |
| Elevated plasma calcitonin | ○ | ○ |
| Pheochromocytomas | ○ | ○ |
| Hypertension | ○ | ○ |
| Head/neck paraganglioma | ○ | ○ |
| Elevated catecholamine levels | ○ | ○ |
| Hyperparathyroidism | ○ | ○ |
| Hypercalciuria | ○ | ○ |
| Kidney stones | ○ | ○ |
| Parathyroid adenomas | ○ | ○ |
| Pruritic cutaneous lichen amyloidosis | ○ | ○ |

#### Presence of MEN2B Symptoms

| | Yes | No |
|---|---|---|
| Mucosal neuromas on tongue, palate, pharynx | ○ | ○ |
| Prominent lips, submucosal nodules | ○ | ○ |
| Neuromas of eyelids | ○ | ○ |
| Thickened corneal nerves | ○ | ○ |
| Ganglioneuromatosis of GI tract | ○ | ○ |
| Abdominal distention, megacolon, constipation | ○ | ○ |

| | | |
|---|---|---|
| Marfanoid habitus | ○ | ○ |
| Kyphoscoliosis | ○ | ○ |
| Lordosis | ○ | ○ |
| Joint laxity | ○ | ○ |
| Decreased subcutaneous fat | ○ | ○ |
| Muscle wasting and weakness | ○ | ○ |

#### Presence of Hirschsprung Symptoms

| | Yes | No |
|---|---|---|
| Enlargement of bowel | ○ | ○ |
| Constipation or obstipation | ○ | ○ |
| Vomiting | ○ | ○ |

#### Presence of Other Sporadic Tumors

| | Yes | No |
|---|---|---|
| Papillary thyroid carcinoma | ○ | ○ |
| Lung adenocarcinoma | ○ | ○ |
| Chronic myelomonocytic leukemia | ○ | ○ |
| Other tumor | ○ | ○ |

Type of tumor ______________________________

#### Labs/Imaging/Procedures

Endocrine Testing
- ☐ Plasma Calcitonin Level
- ☐ Plasma CEA
- ☐ Plasma Free or 24-hour Urine Metanephrines
- ☐ Serum Parathyroid Hormone (PTH) Level
- ☐ Serum Calcium Level
- ☐ Other Endocrine Test

Plasma Calcitonin Results ______________________________

Plasma CEA Results ______________________________

Plasma Free or 24-hour Urine Metanephrines Results ______________________________

Serum PTH Results ______________________________

Serum Calcium Results ______________________________

Other Endocrine Test Results

__________________________________

Imaging

☐ Neck Ultrasound
☐ MRI Abdomen
☐ CT Abdomen
☐ Liver MRI
☐ FDOPA PET
☐ MIBG Scintigraphy
☐ Other Imaging

Neck Ultrasound Results

__________________________________

MRI Abdomen Results

__________________________________

CT Abdomen Results

__________________________________

Liver MRI Results

__________________________________

FDOPA PET Results

__________________________________

MIBG Scintigraphy Results

__________________________________

Other Imaging Results

__________________________________

Hirschsprung Tests

☐ Rectal Suction Biopsy
☐ Barium Enema
☐ Anorectal Manometry

Biopsy Results

__________________________________

Enema Results

__________________________________

Anorectal Manometry Results

__________________________________

| | |
|---|---|
| Procedures | ☐ Thyroidectomy<br>☐ Adrenalectomy<br>☐ Parathyroidectomy<br>☐ Tumor Resection<br>☐ Other Procedure |
| Thyroidectomy Notes | __________________________________ |
| Adrenalectomy Notes | __________________________________ |
| Parathyroidectomy Notes | __________________________________ |
| Tumor resection notes | __________________________________ |
| Other Procedure Notes | __________________________________ |
| Other Comments | __________________________________ |
| Eligible for recontact? | ◯ Yes<br>◯ No |
| Has symptoms of RET conditions? | ◯ No/limited symptoms<br>◯ Moderate symptoms<br>◯ Marked symptoms<br>◯ Not enough information in chart |

### WT1 Chart Review

Record ID ______________________

Variant ______________________

Medical Record Number ______________________

MGB Biobank Number ______________________

#### Demographic Information

Sex
- ◯ Male
- ◯ Female

Age ______________________

Race
- ☐ American Indian or Alaskan Native
- ☐ Asian Indian
- ☐ Black or African American
- ☐ East Asian
- ☐ Native Hawaiian or Pacific Islander
- ☐ Other
- ☐ White

Ethnicity
- ◯ Hispanic
- ◯ Not Hispanic

Dead/Alive
- ◯ Dead
- ◯ Alive

Cause of Death ______________________

Death related to variant?
- ◯ Yes
- ◯ No

#### Diagnosis

Had genetic testing?
- ◯ Yes
- ◯ No

WT1 Disorder Diagnosis?
- ◯ Yes
- ◯ No

Age at genetic diagnosis ______________________

Type of genetic testing ______________________

Seeing genetics? ◯ Yes ◯ No

Problem list __________________________________

#### Family History

Family history __________________________________

#### Family History Suggestive of WT1 Disorders

| | Yes | No |
|---|---|---|
| Diagnosis of WT1 disorder | ◯ | ◯ |
| Wilms tumor | ◯ | ◯ |
| Kidney or urological cancers | ◯ | ◯ |
| Gonadoblastoma | ◯ | ◯ |
| Congenital nephrotic syndrome | ◯ | ◯ |
| Kidney failure | ◯ | ◯ |
| Chronic kidney disease | ◯ | ◯ |
| Persistent proteinuria | ◯ | ◯ |
| Genitourinary abnormalities | ◯ | ◯ |
| Diaphragmatic hernia | ◯ | ◯ |

#### Specialists Seen

Specialists seen
- ☐ Nephrology
- ☐ Oncology
- ☐ Endocrinology
- ☐ Urology
- ☐ Gynecology
- ☐ Psychology
- ☐ Other Specialists

Last Nephrology A/P __________________________________

Last Oncology A/P __________________________________

Last Endocrinology A/P __________________________________

Last Urology A/P __________________________________

Last Gynecology A/P

______________________________

Last Psychology A/P

______________________________

Other Specialists seen

______________________________

#### Presence of WT1 Disorder Symptoms

| | Yes | No |
|---|---|---|
| Persistent proteinuria | ◯ | ◯ |
| Early-onset SRNS | ◯ | ◯ |
| Hypoalbuminemia | ◯ | ◯ |
| Edema | ◯ | ◯ |
| Hyperlipidemia | ◯ | ◯ |
| Chronic kidney disease | ◯ | ◯ |
| Congenital nephrotic syndrome | ◯ | ◯ |
| Wilms tumor | ◯ | ◯ |
| Testicular development | ◯ | ◯ |
| External genitalia abnormalities | ◯ | ◯ |
| Complete gonadal dysgenesis | ◯ | ◯ |
| Internal genitalia abnormalities | ◯ | ◯ |
| Streak gonad | ◯ | ◯ |
| Dysgenetic testes | ◯ | ◯ |
| Gonadoblastoma | ◯ | ◯ |
| Duplex kidney | ◯ | ◯ |
| Horseshoe kidney | ◯ | ◯ |
| Kidney malrotation | ◯ | ◯ |
| Vesico-urinary reflux | ◯ | ◯ |
| Pelviureteric junction | ◯ | ◯ |
| Urogenital sinus | ◯ | ◯ |
| Diaphragmatic hernia | ◯ | ◯ |

#### Labs/Imaging/Procedures

Blood Testing

- ☐ Urinalysis
- ☐ Serum protein
- ☐ Serum albumin
- ☐ Serum creatinine
- ☐ Serum cholesterol
- ☐ Serum IgG
- ☐ Serum C3
- ☐ Blood pressure
- ☐ BUN
- ☐ Other Test

Urinalysis Results

______________________________

Serum Protein Results

______________________________

Serum Albumin Results

______________________________

Serum Creatinine Results

______________________________

Serum Cholesterol Results

______________________________

Serum IgG Results

______________________________

Serum C3 Results

______________________________

Blood Pressure Results

______________________________

BUN Results

______________________________

Other Test Results

______________________________

Imaging

- ☐ Renal US
- ☐ Pelvic US
- ☐ Abdominal US
- ☐ Chest X-Ray
- ☐ Other Imaging

Renal US Results

______________________________

Pelvic US Results

______________________________

Abdominal US Results

______________________________

Chest X-Ray Results ______________________

Other Imaging Results ______________________

Procedures
- ☐ Kidney Biopsy
- ☐ Gonadal Biopsy
- ☐ Urodynamic Studies
- ☐ Hormonal Studies
- ☐ Removal of Wilms Tumor
- ☐ Nephrectomy
- ☐ Gonadectomy
- ☐ Other Procedure

Kidney Biopsy Results ______________________

Gonadal Biopsy Results ______________________

Urodynamic Studies Results ______________________

Hormonal Studies Results ______________________

Removal of Wilms Tumor Results ______________________

Nephrectomy Results ______________________

Gonadectomy Results ______________________

Other Procedure Results ______________________

Other Comments ______________________

Eligible for recontact?
- ◯ Yes
- ◯ No

Has symptoms of WT1 Disorder?

◯ No/limited symptoms
◯ Moderate symptoms
◯ Marked symptoms
◯ Not enough information in chart
